# STeCC: Smart Testing with Contact Counting Enhances Covid-19 Mitigation by Bluetooth App Based Contact Tracing

**DOI:** 10.1101/2020.03.27.20045237

**Authors:** Hossein Gorji, Markus Arnoldini, David F. Jenny, Alexandre Duc, Wolf-Dietrich Hardt, Patrick Jenny

## Abstract

Covid-19 mitigation commonly involves social distancing. Due to its high economic toll and its impact on personal freedom, we need to ease social distancing and deploy alternative measures, while preventing a second wave of infections. Bluetooth app-based contact tracing has been proposed, focusing on symptomatic cases and isolating their contacts. However, this approach would miss many transmissions by asymptomatic cases. To improve effectiveness of app-based mitigation, we propose to complement contact tracing with Smart Testing relying on Contact Counting (STeCC). STeCC focuses virus RNA testing to people with exceptionally high numbers of contacts. These people are at particularly high risk to become infected (with or without symptoms) and transmit the virus. Mathematical modeling shows that a mitigation strategy combining STeCC and contact tracing in one app will be more efficient than contact tracing and works when ≈50% (instead of ≥60%) of the total population participate. Similarly, it requires 50-100 fold less tests than randomized virus testing alone. These gains in efficiency may be critical for success. STeCC could be integrated in the current Bluetooth tracing apps. Thus, STeCC is technically feasible and can reduce the pandemic’s reproduction number by 2.4-fold (e.g. from R_0_=2.4 to R_eff_=1) with realistic test numbers (≈166 per 100’000 people per day), when a realistic fraction of the population would use the app (i.e. ≈50% in total population). Thereby, STeCC efficiently complements the portfolio of mitigation strategies, which allow easing social distancing without compromising public health.

## Introduction

The Covid-19 pandemic has evaded initial containment measures. Current responses have therefore shifted towards mitigating the effects. However, proven vaccines and therapies are lacking and the current capacity for detecting the virus via its genomic RNA is limited^1^. Thus, mitigation in many countries relies on a broad portfolio that includes not only hygiene measures, but also physical distancing (termed social distancing, hereafter) to reduce transmission, diagnosing virus (SARS-CoV2) infections in infected people showing mild to severe symptoms and tracing of their recent contacts. In combination, these measures slow down pandemic spread and help to avoid overburdening healthcare systems by easing the demand for intensive care. However, this strategy has two major shortcomings. First, it leaves many infected people with mild or no symptoms undetected^2^, and therefore renders them more likely to infect others. Second, as social distancing measures limit non-essential business, it imposes severe economic consequences, which worsen over time. A broad social-distancing-based approach is therefore not sustainable, but mitigation measures need to stay in place until effective therapies or vaccines become available to avoid a second wave of virus spread. As these options are still months to years away, we need to consider alternative mitigation strategies.

Smartphone apps for Bluetooth-based contact tracing, such as the Swiss D3-PT, the British NHSX or the European PEPP-PT project can help to identify individuals that have recently had an infection-relevant contact (i.e. one that confers a risk of transmission) with known Covid-19 cases and might therefore have been infected. Self-quarantine of these contacts can help to mitigate the pandemic^3,4^. However, the efficiency of this approach is limited by app-related features (e.g. the percentage of users in the population, efficiency in identifying infected people, efficiency in identifying relevant contacts), and by uncertainties about important epidemiological characteristics of the virus (e.g. the virus’ epidemic doubling time, the fraction of pre-symptomatic and asymptomatic cases, and their infectiousness). Infected people who are not showing symptoms are particularly important: for example, the study of the COVID-19 outbreak in the municipality of Vo’, Italy conducted by Lavezzo et al.^5^, performed two virus tests two weeks apart, and observed that 43.2% of infected people were asymptomatic and that their viral loads were as high as symptomatic cases. Contact tracing of symptomatic cases by Bluetooth apps or traditional methods would have missed these cases and failed to detect and quarantine their contacts. Infectiousness of asymptomatic cases has been estimated to be between 10% and 100% of symptomatic cases^4-6^ (based on differences in viral load), and the fraction of asymptomatic cases has been reported to be between 11.5% and 43.2% of all infected cases^6-8^. Asymptomatic cases are therefore a particular problem, as app-based contact tracing detects such cases insufficiently and thereby reduces the efficiency of this approach (**Fig. S5**). Therefore, we have looked for ways to improve the efficiency of this approach. We focused on testing for virus RNA, a marker for active infections with or without symptoms (**Fig. 1**, highlighted in dark or light blue).

**Figure 1.**
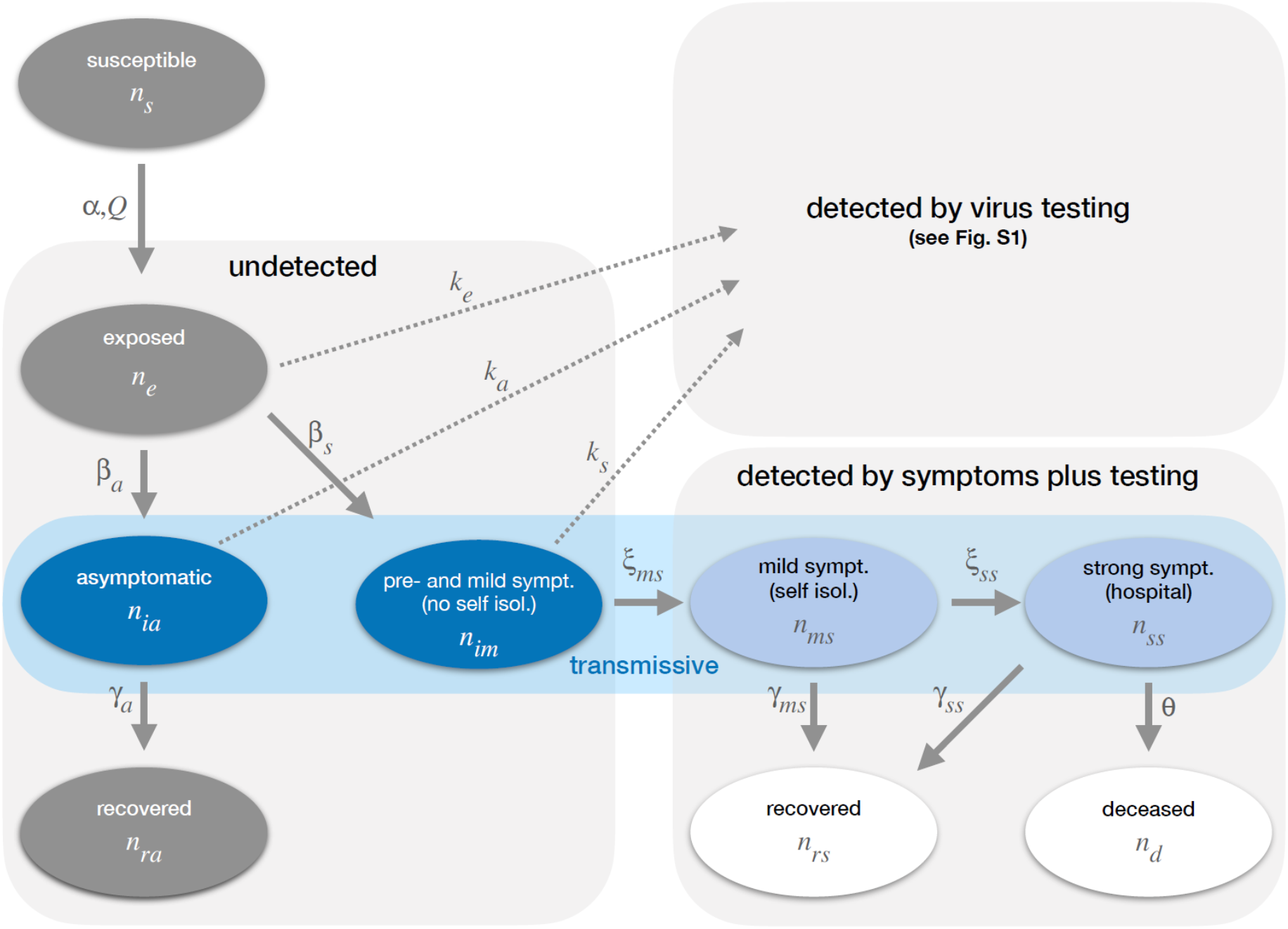
Graphical illustration of the modeling approach showing the dependencies within the system describing the dynamics of the susceptible, undetected and detected infected populations. It is crucial that the model distinguishes between individuals detected by symptoms (light blue), and those detected by virus testing.

## Results

First, we asked how well mass testing random samples of the population could work. In theory, this can be achieved by testing large fractions of the population at regular intervals, and isolating people who test positive. Using a deterministic modeling approach that explicitly considers infected cases with and without symptoms, we can estimate how many tests per 100’000 people per day would suffice to achieve the same effect as current social distancing measures which have reduced the basic reproduction number of the pandemic R_0_ from 2.4-3 to an effective reproduction number R_eff_ of less than 1, e.g. in Germany or Switzerland (^9,10^; for technical details, see **supplementary text, table S2**). We will first present our findings and then discuss how testing could be realistically implemented, i.e. by combining it with app-based technologies.

To estimate how mitigation strategies affect R_eff_, we use a mathematical model that employs a set of ordinary differential equations to describe the dynamics of the infection of a susceptible population (**Fig. 1, supplementary text** section 1, **Fig. S1, table S1**). To obtain realistic simulations, we parametrize our model using published data (^6,11-16^; **table S2; supplementary text** section 2). Infection of susceptible people will lead to a latency phase (**Fig. 1**, exposed). The exposed will later become transmissive, and either remain asymptomatic and recover or become pre-symptomatic and later develop mild symptoms (**Fig. 1**, dark blue; see parameters in **table S2**). These two transmissive groups do not know that they are infected, and remain unidentified in current mitigation approaches. Some people with mild symptoms of disease will self-isolate (**Fig. 1**, light blue). Due to self-isolation, they will have a reduced probability to infect others (we assume a 90% reduction). Infected people with severe symptoms are hospitalized, immediately isolated under strict quarantine and do not infect others (**Fig. 1**, light blue). The same applies for anyone else who is tested virus-positive. In addition, we consider the effect of overloading the health care system. After all intensive care units (ICUs) are occupied, additional cases requiring intensive care will suffer an elevated death rate (**Fig. 1; supplementary text**, section 2). As our model consists of a set of ordinary differential equations, which become linear in the early stages of the pandemic (when nearly the whole population still is susceptible), we can now analytically test how particular mitigation strategies affect R_eff_ (**supplementary text**, sections 3, 4 and 5).

In order to compare different mitigation strategies to a baseline, we first describe what the model predicts if we omit any mitigation. Here, we assume a value for R_0_ (the basic reproduction number in the unmitigated case) that is in the mid-range of published estimates, i.e. that one infected person infects 2.4 others in average (R_0_=2.4; **Fig. 2A;** alternative R_0_ values are tested in **Fig. S2A**,**B**). Under these circumstances, 87% will either recover or die from the disease within ≈250 days, which compares well to the 81% predicted by Ferguson et al.^6^ for UK and US populations in the absence of mitigation plans. We also predict more than 4% of the population to be killed by the virus, and ICUs to be at maximum capacity for ≈150 days of that year. For the unmitigated case these numbers are going to change only by 2-fold or less, if alternative plausible values are assumed for R_0_ (1.9 or 2.9; **Fig. S2A**,**B**) or for the availability of ICU beds (**Fig. S2C**). Thus, our basic predictions are robust, even if input parameters for the virus infection dynamics (**table S2**) might be subject to change when more precise parameters became available. Please note that **Fig. 2A** depicts the worst-case scenario, which can be improved by mitigation.

**Figure 2.**
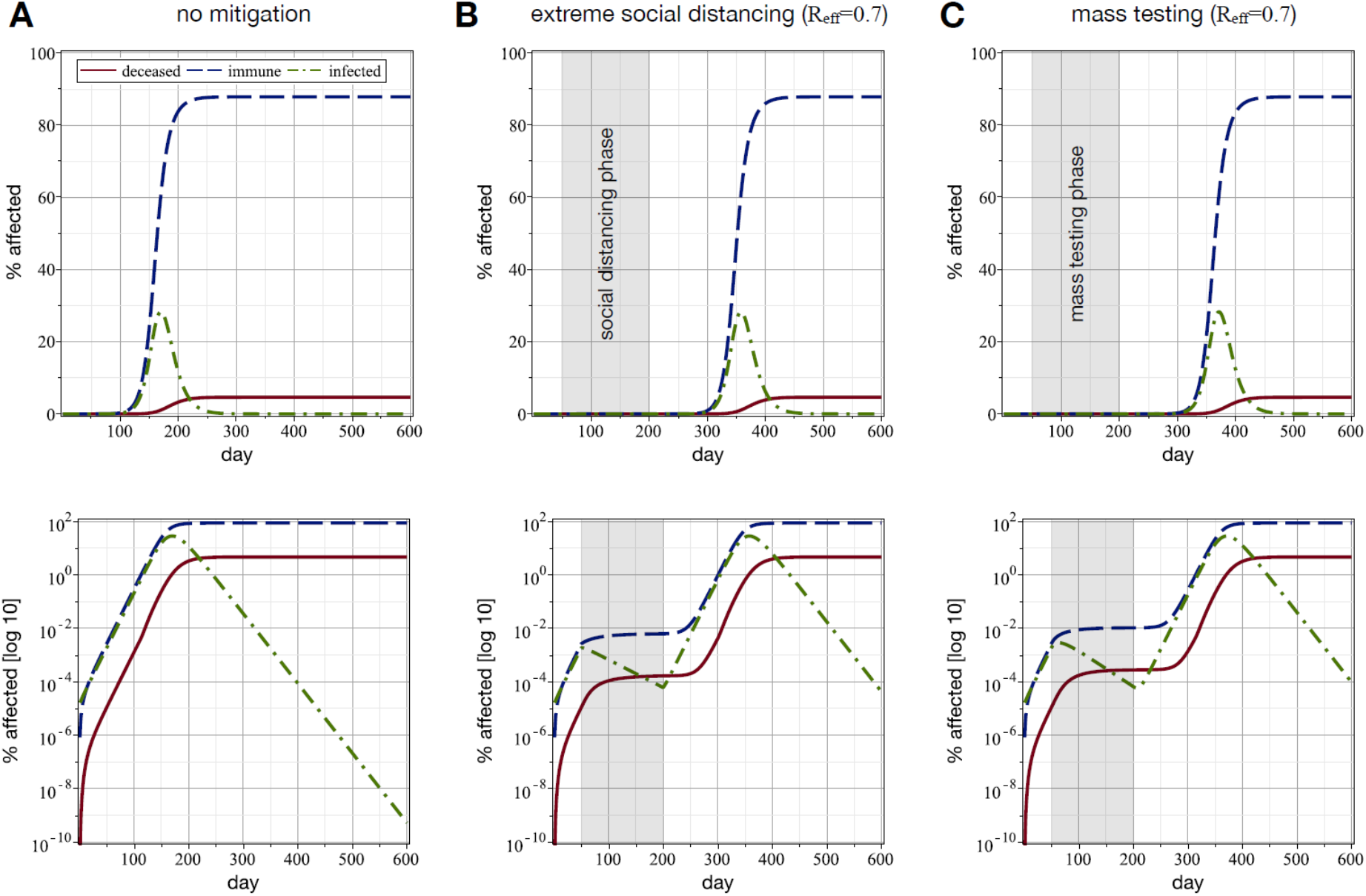
Social distancing and extensive mass testing alone have qualitatively equal mitigating effects. We used the model described in the **supplementary text**. (A) Model outcome if no mitigation strategies are in place (R_0_=2.4; see **table S3** for additional input parameters). (B) Model outcome if extreme social distancing (71% lower infection rate, leading to R_eff_=0.7; **table S2**) is in place between days 50 and 200 of the outbreak. After day 200, social distancing is discontinued. (C) Model outcome if mass testing with isolation of detected cases is applied between days 50 and 200 of the outbreak (as shown in A). The shown effect is achieved if 37’300 people per 100’000 are tested every day (5% false negative rate; test speed = 1 day). **Tables S2 and S3** show all other model parameters. Testing is discontinued after day 200. Dashed blue lines represent recovered plus infected plus deceased, dashed-dotted green lines infected, and solid red lines deceased people. Lower panels show the same data as upper panels, but with a log_10_-scale for Y-axes.

We then compare the effects of extreme social distancing with those of extensive mass testing. Extreme social distancing is imposed for a period of 150 days (day 50-200 of the pandemic), reducing R_eff_ to 0.7 (**table S1**). This is achieved if the rate of infection of susceptible people is reduced by 71% (**Fig. 2B**; data for R_eff_=1, as recently achieved or surpassed in Switzerland^10^ and other European countries, is shown in **Fig. S4A**). In line with current observations, extreme social distancing dramatically decreases the fraction of people experiencing an infection and reduces the number of deaths in our model (<0.0001% in **Fig. 2B**), at least if social distancing is initiated early on (e.g. by day 50) and strictly adhered to for months. The fraction of the infected population declines during extreme social distancing. This is in line with other modeling studies^3,4^. However, once these measures are abandoned, the infection starts spreading again, leading to a similar death toll and ICU overloading as in the case without mitigation (**Fig. 2B**, days 200-600).

Next, we analyze a mitigation strategy that is based on mass testing alone (**supplementary text** section 3). We would test random samples of the population with concomitant isolation of detected cases. The test is assumed to take one day to process, to yield 5% false negative results (discussed below), which is technically reasonable, and to result in immediate quarantine of the virus-positive cases. **Figure S3A** shows the factor by which R_eff_ changes as a function of number of tests and for different test processing times; the same is shown in **Fig. S3B** for a higher false negative rate. Again, we assume that the mitigation strategy is implemented during days 50-200 of the pandemic. Applying 37’300 tests per 100’000 people per day yields the same outcome as extreme social distancing (R_eff_=0.7; **Fig. 2C**, compare to **Fig. 2B; supplementary text** section 3 for details). Reducing the mass testing to 12’600 tests per 100’000 people per day is sufficient to keep the number of infections constant (R_eff_=1, **Fig. S4B**) and yields equivalent results as a moderate form of social distancing (**Fig. S4A**). Both interventions can dramatically slow the epidemiological dynamics, but similarly to the previously discussed social distancing, the number of infections and deaths will start to rise again after the testing regime is abandoned (**Fig. 2C**, days 200-600).

It is easy to see that both, mass testing and social distancing, can have qualitatively identical effects on the epidemiological dynamic, but they work differently. Social distancing decreases the overall infection rate by reducing the freedom of movement for all, whereas mass testing will allow us to limit the isolation to the infected fraction of the population that would transmit the virus. The latter would have the important advantage of inflicting much smaller burdens on the economy than social distancing, as far smaller fractions of the population need to be isolated. However, the number of tests we predict to be necessary to reach R_eff_≤1 (12’600 tests per 100’000 people per day; **Fig. S4B**) is clearly unrealistic for now, and >50-fold above the current testing capacities even of countries with highly developed and well-funded healthcare systems. Optimizing the test specificity or the test speed changes the required number of tests by merely <2-fold (**Fig. S3**). Thus, mass testing alone is not an option.

Next, we analyzed mitigation strategies combining virus testing with other measures. This should reduce the number of tests needed to achieve R_eff_ ≤1. First, we tested a mitigation strategy combining mild social distancing, which reduces the infection rate by 33% (this would yield R_eff_=1.6, if applied alone; **table S1**), and mass testing (assumed test speed = 1 day, 5% false negatives). This may approximate a scenario where mass testing is applied to balance the effects of easing current social distancing measures. Using this combined strategy, we would require 4’500 tests per 100’000 people per day to reach R_eff_=1 (**Fig. 3A**). However, this still exceeds the current test capacities.

**Figure 3.**
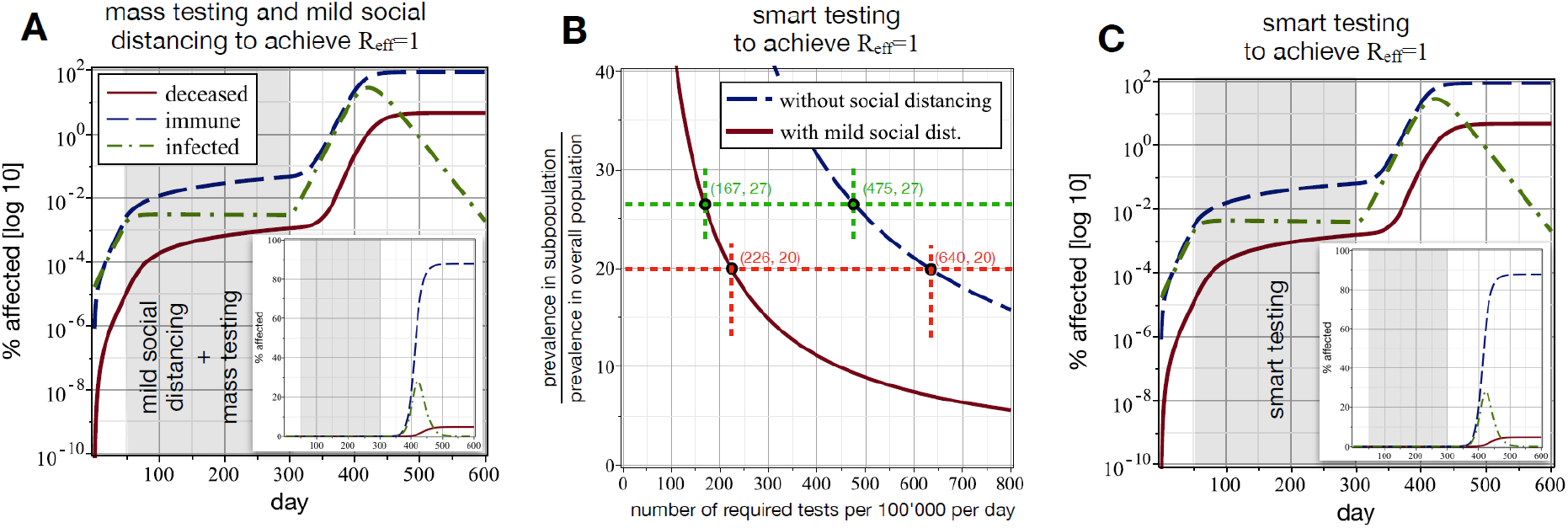
Effective combinations of mass testing with other mitigation strategies can be achieved with realistic numbers of tests. (**A**) A combination of mild social distancing (33% reduction in infection rate; R_eff_=1.6 if it were applied alone) and mass testing (4’500 tests per 100’000 per day; false negative rate = 5%; test speed = 1 day) during days 50-300 can reduce R_eff_ to 1. (**B**) Smart selection of the tested subpopulation reduces the required test numbers, if additional information allows identifying a subpopulation with higher than average infection prevalence (details, below). The Y-axis shows the factor by which the infection prevalence increases in the tested subpopulation. This will reduce the required number of tests (X-axis). Blue dashed line: smart testing alone is used to achieve R_eff_=1 (if R_0_=2.4 for the case without mitigation); red solid line: smart testing combined with mild social distancing (33% reduction in infection rate by social distancing). Green and red points illustrate examples discussed, below. (**C**) The same effect as in A can be achieved with smart testing, assuming that we can find a subpopulation with a 27-fold increased infection prevalence. Here, we need 475 tests per 100’000 per day to reach R_eff_=1 (green dots in panel B). (A,C): Dashed blue lines: recovered plus infected plus deceased; dashed-dotted green lines: infected; solid red lines: deceased people. Inserts show the same data, but with a linear scale for Y-axes. All presented cases assume false negative rate = 5%; test speed = 1 day.

An additional mitigation measure is serological testing to identify the subpopulation that has already recovered from the disease and is now immune^17^. This could be combined with mass testing to reduce the number of tests by removing the immune subpopulation from the pool of candidates to be tested. However, we are still at a stage of this pandemic where the fraction of immune people is likely quite small (see **Fig. 2A**). Therefore, serological testing could only minimally reduce the number of needed tests per 100’000 people per day. In no way are we implying, however, that serological testing is not useful: especially in the case of healthcare workers, and other essential personnel that is in close contact with the public, it is of great value to know who is immune to the disease. Also, serological testing could complement mitigation at later phases of such an epidemic when larger fractions of the population have become immune (**Fig. 2A**; >10% immune or deceased beyond day 140).

Finally, we assessed how virus testing could be combined with Bluetooth-based tracing applications (e.g. D3-PT, NHSX or PEPP-PT), which are designed to identify individuals who had recent contacts with infected people and might therefore be infected, too. Contact tracing alerts the detected contacts to encourage isolation or virus testing. However, this strategy leaves room for improvement, as it misses most transmission events by asymptomatically infected cases, pre-symptomatic cases and cases with mild symptoms that remained undetected (**Fig. 1**, dark blue; **Fig. S5, supplementary text section 4**). We explored a complementary approach that uses information from tracing apps to develop a smart testing strategy. In theory, smart testing would use additional information to focus virus tests on subpopulations having a higher prevalence than the overall population. This would include asymptomatic cases, pre-symptomatic cases, symptomatic cases, and their contacts. We will first analyze the theoretical principle and then discuss a practical implementation.

For smart testing to work, two crucial requirements must be fulfilled. The prevalence in the tested subpopulation has to be high enough, so that a limited number of tests will suffice to detect the necessary number of infected people. Furthermore, the traced subpopulation featuring the desired prevalence has to be large enough. We describe the detailed derivation of the corresponding mathematical expressions in the **supplementary text** section 5. Smart testing alone can substantially reduce the number of virus tests needed to achieve R_eff_=1 (**Fig. 3B**, dashed blue line). In fact, if we assume that smart testing identifies a subpopulation with 27-fold higher prevalence than the overall population, we can achieve R_eff_=1 with 475 tests per 100’000 per day (**Fig. 3B**, dashed blue line, green dot) and thus keep the number of infections constant as long as smart testing is maintained (**Fig. 3C**, days 50-300). Fewer tests would be needed, if we could further increase the virus prevalence in the tested subpopulation (or identify people with a higher than average chance to transmit the virus). Alternatively, one could combine smart testing with mild social distancing, e.g. to stop a surge of incidence when strong distancing measures are eased. For example, if social distancing would reduce the infection rate by merely 33%, we would only need 167 tests per 100’000 per day to reach R_eff_=1 (**Fig. 3B**, red line, green dot). Sufficient testing capacity for this combined strategy would already be available today in several European countries. We conclude that smart testing could work: it could achieve the same result as mass testing (**Fig. S4B**), but with much fewer tests (**Fig. 3B**).

Can we use Bluetooth-based tracing apps for identifying high prevalence subpopulations? A first attempt at achieving this would be an adaptation of app-based contact tracing to identify individuals with recent infection-relevant contact to detected Covid-19 cases^4^. This contact subpopulation should have a higher prevalence than the overall population. However, such a “trace+test” approach is still limited by its focus on contacts of symptomatic individuals (while contacts to asymptomatic cases are missed), as well as the virus’ infection and transmission dynamics (for details, see the **supplementary text** section 6). Contact tracing as a selection procedure for smart testing will therefore suffer from the same limitations as contact tracing itself.

As many asymptomatic SARS-CoV2 carriers contribute to the transmission, we reasoned that smart testing should cover symptomatic and asymptomatic cases, alike. To solve this problem, we suggest a different strategy for identifying high-prevalence subpopulations. Importantly, this can be achieved using the same Bluetooth based technology as contact tracing. Our strategy relies on the fact that some individuals will have many more infection-relevant contacts than most others^18^. For example, with realistic scale-free network model assumptions, ≈1% of the population with the most infection-relevant contacts (at least 10 times more than the average) would make the difference between R_eff_=1 and R_0_=2.4 (see **supplementary text** section 6). These individuals can be identified by counting of infection-relevant contacts (i.e. contacts which could facilitate transmission), regardless, if contacts were infected or not. Their sheer number of contacts makes high-contact individuals much more likely than others to become infected and to transmit the virus, and they are known to be highly important for epidemiological dynamics (^19^, referred to as super-spreaders). To test if smart testing with contact counting (STeCC; **Fig. 4A**) works in realistic scenarios, we used a scale free network model, which can account for the heterogeneity of the number of contacts in a population (**supplementary text** section 6). The fraction of app-users in the population will also affect the success of STeCC. We assume that children under age 10 contribute little to transmission in the general population ^20,21^ and would not use smartphones, as suggested by others ^3^. High-risk persons (i.e. >70 years old; 20% of the overall population) would remain shielded in isolation for safety reasons and would therefore not contribute to the infection dynamics. People aged 10-70 (in particular the high-contact subgroup) would use smartphones, and we assessed how their app-usage would affect mitigation.

**Figure 4.**
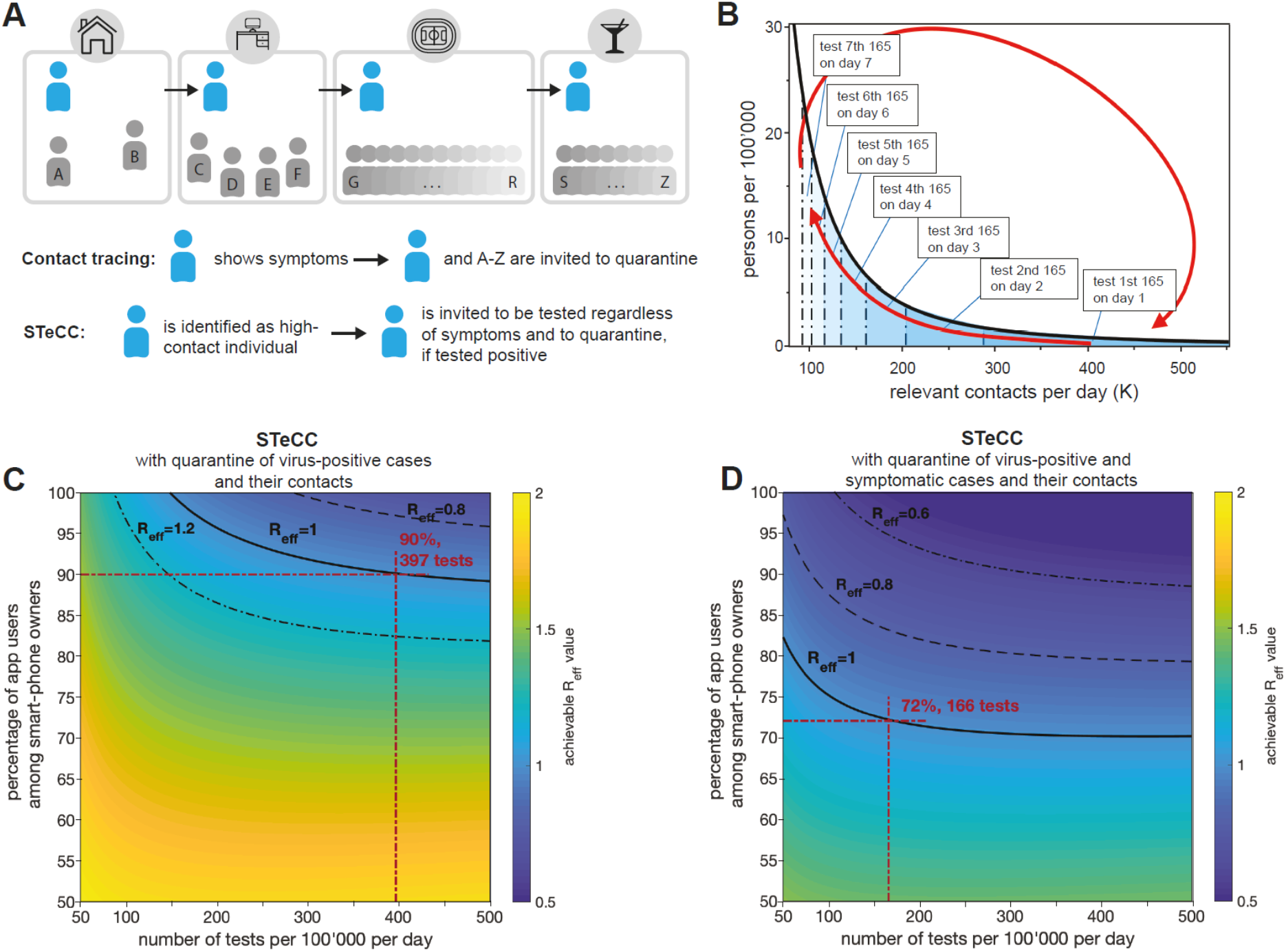
STeCC can help to achieve R_eff_=1 using a realistic number of tests and realistic rates of app-usage. (**A**) STeCC approach vs contact tracing. Contacts of a focal individual are logged using software. Using contact tracing, all contacts would be quarantined if the focal individual shows symptoms. Using STeCC, the focal individual is tested for infection if their contacts exceed a threshold number (e.g., if they visit a sporting event and a bar, in addition to being at home and at work), and quarantined if tested positive. (**B**) Progressive test cycling approach. Example for 80% app users among smart-phone owners and the availability of 165 tests per 100’000 people per day. (**C**) Covid-19 mitigation by STeCC alone with quarantine of contacts of virus-positive cases. Graph showing how the number of tests per 100’000 per day relates to the percentage of app users. We assume R_0_=2.4, test speed of 1 day, immediate notification and quarantine, false negative ratio 5%, asymptomatic cases (1/3 of all) are 50% as infective as symptomatic cases. Solid line: values to achieve R_eff_=1; dashed line: values to achieve R_eff_=0.8; dashed dotted line: values to achieve R_eff_=1.2. (**D**) Covid-19 mitigation by combining STeCC with app-based contact tracing. Graph showing how the number of tests per 100’000 per day relates to the percentage of app users. We assume R_0_=2.4, test speed of 1 day, immediate notification and quarantine, false negative ratio 5%, asymptomatic (1/3 of all) are 50% as infective as symptomatic cases. Solid line: values to achieve R_eff_=1; dashed line: values to achieve R_eff_=0.8; dashed dotted line: values to achieve R_eff_=0.6. The coloring indicates the achievable R_eff_ with the respective mitigation strategy in place (see **supplementary text**, sections 6).

Then, we tested three different scenarios how STeCC could be implemented to reach R_eff_=1. We used a progressive testing cycle, as testing the same high-contact individuals every day would seem unrealistic. On the first day, based on the number of available tests, we would identify the optimal subgroup with the highest frequency of infection-relevant contacts, and invite them to be tested (**Fig. 4B**; dark blue; **supplementary text** section 6). During the next days, we would repeat the same procedure, but exclude the groups previously identified. After 7 days, we would begin the cycle again and identify the optimal high-contact group from the entire susceptible population. Using this testing cycle, we observed that STeCC alone could reduce R_eff_, but cannot reach R_eff_=1 with realistic numbers of app-users and tests per day (**Fig. S6**). Next, we assessed a STeCC strategy, which would refer two groups of people to quarantine, i.e. the high-contact individuals that tested positive and their recent contacts. This strategy would reach R_eff_=1, if 90% of smartphone users would use the app (63% of the overall population) and ≈397 tests per 100’000 people per day were carried out (**Fig. 4C**, red lines). Finally, we tested a combination of the latter STeCC approach with Bluetooth app-based contact tracing (similar to ^4^), which may be possible using the same data. This would refer three groups of people to quarantine, i.e. the high-contact individuals testing positive, their recent contacts and the contacts of symptomatic individuals. If 72% of smartphone users would use the app (≈50% of the overall population) we would need ≈166 tests per 100’000 people per day to reach R_eff_=1 (**Fig. 4D**, red lines). This testing capacity is currently available in several European countries, like Switzerland. Please note that our example in **Fig. 4D** assumes an R_0_=2.4, if no mitigation was applied. An identical 2.4-fold reduction is achievable for any other basic virus reproduction rate, as long as 72% of smartphone users would use the app and ≈166 tests per 100’000 people per were carried out. We have conducted sensitivity studies to study effects of the false negative rate, network parameters, test processing times and R_0_. In all considered variations, STeCC with contact tracing could achieve R_eff_=1 with reasonable fractions of app-users and test numbers (see **supplementary text** section 6). This strategy is always more efficient that app-based contact tracing or randomized virus testing alone. Thus, STeCC with contact tracing could add to any mitigation policy.

## Discussion

What are the advantages of a STeCC-based mitigation strategy? First, it includes detection and removal of asymptomatic and pre-symptomatic cases and their contacts. This increases the efficiency of app-based approaches. For example, in contrast to contact tracing alone (**Fig. S6**), the combination with STeCC would work when >72% of the smartphone users (≈50% of the overall population) would use the app, or if unfavorable assumptions about the pandemic’s parameters would turn out to be true (**Fig. S8**). This synergy is attributable to the different selection processes of both approaches. Furthermore, STeCC could be implemented within the same apps as developed for contact tracing. In **supplementary text**, section 7, we are suggesting one possible implementation. This could offer additional choices for app-based mitigation, i.e. the parallel use of STeCC and contact tracing, which is particularly powerful (**Fig. 4D**). As STeCC focusses on a small high-contact group, it works with relatively small numbers of tests and would only quarantine those that are virus positive (and their contacts, e.g. **Fig. 4C**,**D**). It is realistic given the available test capacities in several countries and would affect smaller fractions of the population than other mitigation strategies (i.e. contact tracing or social distancing). Thereby, STeCC will enhance the impact of Bluetooth-based tracing applications. Due to its key function in virus transmission, the identified high-contact subgroup would also be a promising priority for vaccination, once limited supplies of a vaccine become available.

What are the limitations of STeCC? First, we cannot exclude that some assumptions used in our model may be too optimistic or that more precise information might be derived later from alternative, more detailed simulation approaches. The percentage and infectiousness of asymptomatic cases, and the distribution of contacts in the population would have the biggest impact. However, modeling the impact with less favorable parameters verified that STeCC would still provide substantial benefits (**Fig. S7, Fig. S8**). Second, false positive virus tests might dilute the ability to detect positive cases if the virus prevalence is very low. Third, contact counting has not been a focus during the development of Bluetooth-based proximity testing applications. Thus, small, but doable adaptations might be needed, in order to enable efficient detection and notification of high-contact individuals (**supplementary text**, section 7).

Strict social distancing has been successful in achieving R_eff_<1 in many countries, but at a high economical and societal cost. Easing of these measures is presently being discussed or implemented. However, if a large fraction of the population has remained susceptible, a second wave of disease is bound to occur in the absence of effective alternative mitigation strategies. We suggest using a combination of contact tracing and STeCC, as a simple mitigation approach which relies on identifying high-contact individuals, testing them for infection, and quarantining positive cases. STeCC requires the same information as contact tracing (a list of unique contacts in a given period of time), and can be implemented using the same information as the contact tracing apps that are currently being developed or already in use. STeCC would be achievable using a number of tests that is realistic today in several European countries, and can be achieved in many others with appropriate efforts.

STeCC adds to the portfolio of mitigation strategies for the Covid-19 pandemic, and complements approaches like classic contact tracing, hygiene measures, randomized testing of cohorts of interest, or other surveillance tools. It could be deployed quickly in countries with sufficient testing capacities like Switzerland (capacity ≈230 tests per 100’000 per day). A combination with contact tracing might be particularly powerful. Our study provides quantitative estimates for the number of tests needed by starting out with realistic assumptions about the relevant parameters, like number of app users. Once STeCC is applied, one can adjust the strategy flexibly in order to ensure the desired performance. STeCC offers a realistic approach to help relaxing broad social distancing policies in the near future without compromising health, while at the same time providing public health officials with much needed actionable information on the success of their interventions. This will be an important prerequisite for reclaiming our normal public life and initiating economic recovery.

## Materials and Methods

The dynamic model was implemented with Maple 2018. The calculations for mass testing, contact tracing and smart testing were implemented with MATLAB and the Statistics Toolbox Release 2018b. The corresponding codes are available on GitHub via https://github.com/gorjih2/STeCC_preliminary.

## Data Availability

All data used for preparation of this manuscript is publicly available.

## Acknowledgements

The authors are very thankful to Dario Ackermann, who created the website with the simulation tool based on the model presented in this paper. The authors would like to thank Emma Slack, Erik Bakkeren and Noemi Santamaria for helpful comments on the manuscript and Maxime Augier for constructive feedback on the technical implementation of STeCC. HG acknowledges funding from the Swiss National Science foundation (grant number 174060).

## Supplementary Materials

1. Supplementary Text describing the mathematical modeling approaches.

