## Supplementary material for "STeCC: Smart Testing with Contact Counting Enhances Covid-19 Mitigation by Bluetooth App Based Contact Tracing"

### 1. Model

A model is proposed to compute the numbers of infected people whose infection has not (yet) been detected and the numbers of infected persons with a detected infection ($n_i^{undet}$ and $n_i^{det}$), respectively (Fig. 1). Note that detected here refers to persons being isolated, which comprises not only those being tested positive, but also those who feel strong symptoms and thus stay in self-quarantine. It is further important to notice that in the case of SARS-CoV2, the undetected infected people are main contributors to the spread of the pandemic [15]. The exact definitions of detected and undetected, as well as those of all other variables and model parameters are found in Table S1. Furthermore, we compute the number of fatalities ($n_d$) and the number of people who recovered after a detected or an undetected infection. Importantly, we assume that these people will have developed protective immunity and we assume that they cannot be infected again in the considered time frame. The initial susceptible population $n_s^0$ is naive (i.e. it lacks immunity against the infection) and $n_s$ is the number of persons who are susceptible at a given time $t$. In our model we assume that the virus is mainly passed on by undetected asymptomatic and mild symptomatic persons; the detected population with mild symptoms transmits at a much lower rate (because of self-isolation, hygiene precautions in hospitals and/or quarantine). While the model does not consider age dependency, it accounts for higher mortality rates due to temporary shortage of intensive care units. The graph in Fig. 1 (main part) shows the dynamic dependencies. The Covid-19 specific parameters have to be estimated from the available data; their values are listed below. It should be noted, that the implementation of our model allows updating our current estimates with more precise values, as new data come in.

Initially, the entire population is susceptible and can get infected. Infected persons first get exposed and are not infectious before the latency time has passed. Then, they either become asymptomatic or mild symptomatic. Asymptomatic persons eventually recover without symptoms, while the others develop symptoms approximately half a day after the end of the latency time. We assume that persons with mild symptoms isolate themselves approximately one day after onset of symptoms and then either recover or become strong symptomatic, which requires hospitalization. Hospitalized individuals either recover or die. Once $n_s$ becomes smaller, which happens quickly without any measures, the infection rate slows down by a factor of $n_s/n_s^0$. This mechanism of slowing down spread of the epidemic due to a shrinking susceptible population is equivalent to herd immunity. It is crucial for the system dynamics that detected persons are isolated (either by self-isolation at home, by hygienic isolation in hospital setting or in other care facilities, or by organized isolation programs for detected infected people, e.g. in hotel rooms) and thus participate at a much smaller rate or not at all in spreading the disease. We assume that these detected infected people have a 10-fold lower likelihood of infecting others than undetected infected people. All this leads to a dynamic system, which is governed

| terminology | meaning |
|---|---|
| susceptible | persons of the considered population who are susceptible and thus can potentially get infected |
| exposed | infected persons; can not yet transmit the virus |
| asymptomatic | infected persons without symptoms; can transmit the virus |
| pre- and mild sympt. (no self isol.) | infected persons with no or mild symptoms; infectious, but not isolated |
| mild sympt. (self isol.) | infected persons with mild symptoms; infectious and isolated |
| strong symptomatic | infected persons with strong symptoms and thus hospitalized; isolated |
| deceased | persons who died |
| recovered | persons who recovered |
| detected | isolated either after positive testing or after falling ill |
| undetected | persons who are either exposed, asymptomatic or mild symptomatic, but were never contained |
| transmissive | persons who are either asymptomatic or symptomatic |
| extreme social distancing | $\mathcal{R}_{\text{eff}} = 0.7$, if no other mitigation measures are applied; the infection rate is reduced by 71% |
| moderate social distancing | $\mathcal{R}_{\text{eff}} = 1.0$, if no other mitigation measures are applied; the infection rate is reduced by 58% |
| mild social distancing | $\mathcal{R}_{\text{eff}} = 1.6$, if no other mitigation measures are applied; the infection rate is reduced by 33% |
| **variable** | |
| $n_s$ and $n_s^0$ | numbers of susceptible and initially susceptible persons, respectively |
| $n_e$, $\tilde{n}_e$ and $n_e^{tot}$ | numbers of exposed persons; not tested, tested and in total, respectively |
| $n_{ia}$, $\tilde{n}_{ia}$ and $n_{ia}^{tot}$ | numbers of asymptomatic persons; not tested, tested and in total, respectively |
| $n_{im}$, $\tilde{n}_{im}$ and $n_{im}^{tot}$ | numbers of persons with mild symptoms during first day; not tested, tested and in total, respectively |
| $n_{ms}$, $\tilde{n}_{ms}$ and $n_{ms}^{tot}$ | numbers of persons with mild symptoms after first day; not tested, tested and in total, respectively |
| $n_{ss}$, $\tilde{n}_{ss}$ and $n_{ss}^{tot}$ | numbers of persons with strong symptoms; not tested, tested and in total, respectively |
| $n_d$, $\tilde{n}_d$ and $n_d^{tot}$ | numbers of deceased persons; not tested, tested and in total, respectively |
| $n_{ra}$, $\tilde{n}_{ra}$ and $n_{ra}^{tot}$ | numbers of recovered persons who had no symptoms; not tested, tested and in total, respectively |
| $n_{rs}$, $\tilde{n}_{rs}$ and $n_{rs}^{tot}$ | numbers of recovered persons who had symptoms; not tested, tested and in total, respectively |
| $n_i^{undet}$ | undetected infected persons: $n_{ie} + n_{ia} + n_{im}$ |
| $n_i^{det}$ | detected infected persons: $n_{ms} + n_{ss} + \tilde{n}_{ms} + \tilde{n}_{ss} + \tilde{n}_{ie} + \tilde{n}_{ia} + \tilde{n}_{im}$ |
| $\mathcal{K}$, $P_k$ and $\mu$ | number of contacts, its probability density and its average, respectively |
| $\mathcal{K}_0$ | minimum degree of connectivity in the sub-population |
| $(\boldsymbol{pk}_c, \boldsymbol{sk}_c)$ | pair of public and private keys |
| $\mathcal{M}$ | set of IDs |
| **parameter** | |
| $\alpha$ and $A(\tau)$ | rate coefficient for infection and expected infectiousness $\tau$ days after infection, respectively |
| $Q$ | relative infection rate by outside contact (travel) |
| $\epsilon$ | ratio between infection rate of self-quarantined and non-quarantined symptomatic cases |
| $\beta_a$ and $\beta_s$ | rate coefficient for latency of asymptomatic and symptomatic cases, respectively |
| $\theta$ | rate coefficient for mortality of hospitalized cases |
| $\gamma_a$, $\gamma_{ms}$ and $\gamma_{ss}$ | rate coefficients for recovery |
| $\xi_{ms}$ and $\xi_{ss}$ | rate coefficients for successively stronger symptoms |
| $k_e$, $k_a$ and $k_s$ | rate coefficients accounting for testing |
| $\mathcal{R}_0$ | basic reproduction number without mitigation |
| $\mathcal{R}_{\text{eff}}$ | effective reproduction number with mitigation |
| $\mathcal{R}_{\text{eff}}^{symp}$ and $\mathcal{R}_{\text{eff}}^{asym}$ | effective reproduction number of symptomatic and asymptomatic cases, respectively |
| $\mathcal{R}_{\text{eff}}^{wt}$, $\mathcal{R}_{\text{eff}}^{nn}$,$\mathcal{R}_{\text{eff}}^{ST-A}$, $\mathcal{R}_{\text{eff}}^{ST-B}$ and $\mathcal{R}_{\text{eff}}^{ST-C}$ | effective reproduction numbers subject to testing, reshaping the network, STeCC-A, STeCC-B and STeCC-C, respectively |
| $\kappa$ | fraction of basic reproduction number related to symptomatic cases |
| $\zeta$ | percentage of app users among smart-phone owners |
| $\theta^{(sat)}$ and $\theta^{(0)}$ | rate coefficient for mortality of hospitalized cases without and with intensive care units, respectively |
| $\gamma_{ss}^{(sat)}$ and $\gamma_{ss}^{(0)}$ | rate coefficient for recovery of hospitalized cases without and with intensive care units, respectively |
| $N$ | testing interval |
| $N^{-1}$ | testing frequency |
| $\eta$ | fraction of false negative test results |
| $C^{(sat)}/2.5$ | fraction of total population for which intensive care units are available |
| $m$ and $k_c$ | lower and higher cut-off of the connectivity distribution, respectively |
| $\gamma$ | difference between the exponent of the power law model and 2 |
| $T$ | disease transmissibility |
| $\mathcal{E}$, $\mathcal{E}^{ST-A}$, $\mathcal{E}^{ST-B}$ and $\mathcal{E}^{ST-C}$ | efficacy of contact counting, STeCC-A, STeCC B and STeCC C, respectively |
| **operator & function** | |
| $\mathbb{E}(\cdot)$ | expectation |
| $\mathcal{P}_{\mathcal{A}}$ and Prob$\{\mathcal{A}\}$ | probability of the event $\mathcal{A}$ |
| $\text{Enc}_{\boldsymbol{pk}_c}(i, s)$ | cyphertext for ID $i$, random seed $s$ and public key $\boldsymbol{pk}_c$ |
| $\oplus$ | bitwise XOR |
| $\bar{b}$ | NOT operator over $b$ |
| & | AND operator |

**Table S 1.** Terminology and nomenclature of model parameters and variables by the following ordinary differential equations:

$$\dot{n}_s = -\alpha(n_{ia}/2 + n_{im} + \epsilon n_{ms})\frac{n_s}{n_s^0} - Q\frac{n_s}{n_s^0}, \tag{1}$$

$$\dot{n}_e = \alpha(n_{ia}/2 + n_{im} + \epsilon n_{ms})\frac{n_s}{n_s^0} - (\beta_a + \beta_s)n_e + Q\frac{n_s}{n_s^0} - k_e n_e, \tag{2}$$

$$\dot{n}_{ia} = \beta_a n_e - \gamma_a n_{ia} - k_a n_{ia}, \tag{3}$$

$$\dot{n}_{im} = \beta_s n_e - \xi_{ms} n_{im} - k_s n_{im}, \tag{4}$$

$$\dot{n}_{ra} = \gamma_a n_{ia}, \tag{5}$$

$$\dot{n}_{ms} = \xi_{ms} n_{im} - (\gamma_{ms} + \xi_{ss}) n_{ms}, \tag{6}$$

$$\dot{n}_{ss} = \xi_{ss} n_{ms} - (\gamma_{ss} + \theta) n_{ss}, \tag{7}$$

$$\dot{n}_{rs} = \gamma_{ss} n_{ss} + \gamma_{ms} n_{ms} \quad \text{and} \tag{8}$$

$$\dot{n}_d = \theta n_{ss}. \tag{9}$$

Note that $\epsilon \in [0, 1]$ is the transmission reduction factor of the self-isolated individuals. It is assumed that those infected persons who were detected by testing or hospitalized infect much less due to strong isolation and other precautions. Therefore, their effect on the infection rate is neglected here. In order to parametrize the model, besides the rates $\alpha$, $\beta_a$, $\beta_s$, $\gamma_a$, $\gamma_{ms}$, $\gamma_{ss}$, $\xi_{ms}$, $\xi_{ss}$ and $\theta$, also the relative rate $Q$ of infections from outside, i.e., by travel or from the animal world, has to be determined. Further, the initial values of $n_s(t)$, $n_e(t)$, $n_{ia}(t)$, $n_{im}(t)$, $n_{ra}(t)$, $n_{ms}(t)$, $n_{ss}(t)$, $n_{rs}(t)$ and $n_d(t)$ have to be chosen. The variables $\tilde{n}_e(t)$, $\tilde{n}_{ia}(t)$, $\tilde{n}_{im}(t)$, $\tilde{n}_{ra}(t)$, $\tilde{n}_{ms}(t)$, $\tilde{n}_{ss}(t)$, $\tilde{n}_{rs}(t)$ and $\tilde{n}_d(t)$ denote the respective numbers of persons who were tested positive and thus are removed from transmission. The detection rates of exposed ($n_e$), asymptomatic ($n_{ia}$) and mild symptomatic ($n_{im}$) persons due to testing are proportional to $k_e$, $k_a$ and $k_s$, respectively. These individuals are then accounted for by the respective numbers $\tilde{n}_e(t)$, $\tilde{n}_{ia}(t)$ and $\tilde{n}_{im}(t)$; see Fig. S1. Note that the graph in Fig. S1 is very similar as the one in Fig. 1, except that there is no node for susceptible persons (since by definition a susceptible person can not be detected infected) and that there exist sources due to testing (dotted arrows) instead of sinks. To account for the dynamics with testing, the system (1)-(9) has to be augmented by the ordinary differential equations

$$\dot{\tilde{n}}_e = -(\beta_a + \beta_s)\tilde{n}_e + k_e n_e, \tag{10}$$

$$\dot{\tilde{n}}_{ia} = \beta_a \tilde{n}_e - \gamma_a \tilde{n}_{ia} + k_a n_{ia}, \tag{11}$$

$$\dot{\tilde{n}}_{im} = \beta_s \tilde{n}_e - \xi_{ms} \tilde{n}_{im} + k_s n_{im}, \tag{12}$$

$$\dot{\tilde{n}}_{ra} = \gamma_a \tilde{n}_{ia}, \tag{13}$$

$$\dot{\tilde{n}}_{ms} = \xi_{ms} \tilde{n}_{im} - (\gamma_{ms} + \xi_{ss}) \tilde{n}_{ms}, \tag{14}$$

$$\dot{\tilde{n}}_{ss} = \xi_{ss} \tilde{n}_{ms} - (\gamma_{ss} + \theta) \tilde{n}_{ss}, \tag{15}$$

$$\dot{\tilde{n}}_{rs} = \gamma_{ss} \tilde{n}_{ss} + \gamma_{ms} \tilde{n}_{ms} \quad \text{and} \tag{16}$$

$$\dot{\tilde{n}}_d = \theta \tilde{n}_{ss}. \tag{17}$$

The effect of testing is further discussed in Section 3. Next it is described how the parameters can be estimated based on literature data.

### 2. Parameter Estimation

Our devised generalized SEIR model becomes closed once we tune the rate coefficients. These coefficients were computed mainly based on data provided in recently published reports [6, 30]. Before giving the values for transfer

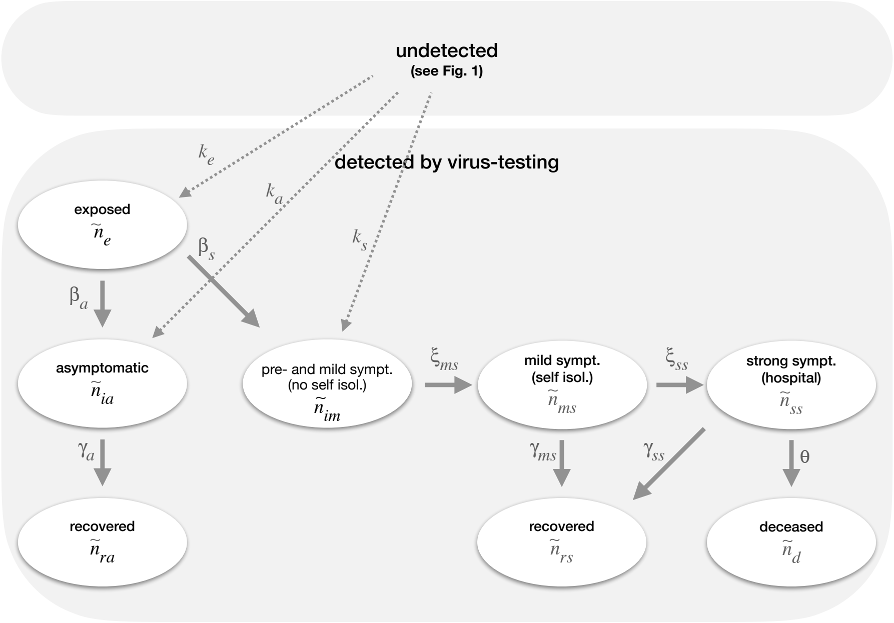


**Fig. S 1.** Graph showing the dependencies of the compartments describing the dynamics of the positively tested people.

rates between different compartments, let us analyze the basic reproduction number $\mathcal{R}_0$ of this virus infection with $Q = 0$ and $n_s(t) \approx n_s^0$. Note that $\mathcal{R}_0$ represents "the expected number of secondary cases produced, in a completely susceptible population, by a typical infective individual" [4]. If $\mathcal{R}_0$ becomes $< 1$, virus spread will decline, and if $\mathcal{R}_0 > 1$, virus spread will increase. To compute $\mathcal{R}_0$, we split the dynamics of the infected population into the infection driven propagation $f$ and the remainder $V$, i.e.,

$$\begin{bmatrix} \dot{n}_e \\ \dot{n}_{ia} \\ \dot{n}_{im} \\ \dot{n}_{ms} \\ \dot{n}_{ss} \end{bmatrix} = \overbrace{\begin{bmatrix} 0 & \alpha/2 & \alpha & \epsilon & 0 \\ 0 & 0 & 0 & 0 & 0 \\ 0 & 0 & 0 & 0 & 0 \\ 0 & 0 & 0 & 0 & 0 \\ 0 & 0 & 0 & 0 & 0 \end{bmatrix}}^{f} \begin{bmatrix} n_e \\ n_{ia} \\ n_{im} \\ n_{ms} \\ n_{ss} \end{bmatrix} - \overbrace{\begin{bmatrix} \beta_a + \beta_s & 0 & 0 & 0 & 0 \\ -\beta_a & \gamma_a & 0 & 0 & 0 \\ -\beta_s & 0 & \xi_{ms} & 0 & 0 \\ 0 & 0 & -\xi_{ms} & \xi_{ss} + \gamma_{ms} & 0 \\ 0 & 0 & 0 & -\xi_{ss} & \theta + \gamma_{ss} \end{bmatrix}}^{V} \begin{bmatrix} n_e \\ n_{ia} \\ n_{im} \\ n_{ms} \\ n_{ss} \end{bmatrix}. \tag{18}$$

Note that testing is not considered here, i.e., $k_e$, $k_a$ and $k_s$ are zero. The $\mathcal{R}_0$ of this system is the spectral radius of $fV^{-1}$, that is,

$$\mathcal{R}_0 \;=\; \rho(fV^{-1}) \;=\; \frac{\alpha\beta_s}{\beta_a + \beta_s}\left[\frac{\beta_a}{2\gamma_a\beta_s} + \frac{1}{\xi_{ms}} + \frac{\epsilon}{\gamma_{ms} + \xi_{ss}}\right]. \tag{19}$$

By inspecting Eq. (19), we observe that we can move towards a stable state (corresponding to $\mathcal{R}_0 \leq 1$) by reducing the infection rate $\alpha$ via mitigation policies such as social distancing. Importantly, as shown later, $\mathcal{R}_0$ can be reduced as well by introducing mass testing, contact tracing, smart testing and subsequent isolation of detected infected individuals.

Next, to clarify our choice of model coefficients, we discuss the rates which appear in transmissive and non-transmissive compartments separately. Finally, the increase of mortality due to lack of intensive care units is modeled.

1. *Transmissive* : We model the incubation time to be log-normally distributed with mean 5.84 (day) and standard deviation 2.98 (day) [28, 20]. In accordance with [6] we take the latency time $x_l$ such that in average it becomes half a day shorter than the incubation time. Similar to the incubation time, we adopt a log-normal distribution for $x_l$ but with mean 5.34 (day) and standard deviation 2.7249 (day). We assume that 1/3 of the cases won't have noticeable symptoms and 2/3 become symptomatic half a day after latency [6]. This leads to $\beta_a = \frac{1}{3}\mathbb{E}(1/x_l) = 0.078$ (1/day), where $\mathbb{E}(\cdot)$ denotes the expectation which gives us the average latency rate. Due to the ratio of 1/3 to 2/3 between asymptomatic and symptomatic cases we get the transfer rate from being exposed to infectious symptomatic as $\beta_s = 2\beta_a = 0.156$ (1/day).

   We suppose that it takes around 1 day from onset of symptoms to self-isolation [6]. Since it takes half a day time delay from becoming infectious to symptomatic, we get $\xi_{ms} = 1/1.5 = 0.6667$ (1/day).

   The average onset to discharge time of clinical cases is around 22 days [30]. We assume that for mild-symptomatic cases the onset to recovery time would be half of this amount, i.e., around 11 days. Therefore the average recovery time from end of the latency period becomes 11.5 days for mild-symptomatic cases. We set the same recovery time for asymptomatic cases which leads to $\gamma_a = 1/11.5 = 0.087$ (1/day).

   A range of values have been suggested for infectiousness of asymptomatic cases; one finds 0.1 in [7], 2/3 in [6] and 1 in [14]. We assume that the asymptomatic cases are 50% less infectious. Furthermore we consider the self quarantined patients to be 90% less infectious, i.e., $\epsilon = 0.1$ is adopted. To compute the infection rate $\alpha$, we assume $\mathcal{R}_0 = 2.4$ [6]. Following Eq. (19), the infection rate becomes $\alpha = 0.6711$ (1/day). Since the basic reproduction number is the most important single parameter of the system, we performed sensitivity studies by changing $\mathcal{R}_0$ [16].

2. *Non-transmissive*: The mean delay time from appearance of symptoms to hospitalization has been reported to be around 11 days [30]. However, 80% of symptomatic cases would not require hospitalization [17]. For those who develop strong symptoms, the delay from self-isolation to hospitalization then becomes $11 - 1 = 10$ days. Hence we get $\xi_{ss} = 0.2 \times 1/10 = 0.02$ (1/day) and $\gamma_{ms} = 0.8 \times 1/10 = 0.08$ (1/day). Note that the latter gives onset to recovery time of 11 days for mild cases consistent with our earlier assumption.

   The average hospital treatment time is 11 days [30, 6]. In case of availability of intensive care units we assume that 20% of hospitalized cases die [17]. Accordingly, we get $\gamma_{ss}^{(0)} = 0.8 \times 1/11 = 0.0727$ (1/day) and $\theta^{(0)} = 0.2 \times 1/11 = 0.0182$ (1/day).

3. *Fatality increase*: We assume that the case fatality ratio increases by two-fold in saturation of the health system. This is justified by noting that the case fatality ratio has increased from approximately 5% in China [2] to roughly 10% in Wuhan while it was the epicentre of the outbreak [22]. By taking this factor into account, and assuming that the average time of hospital treatment remains 11 days, we can compute the death rate of hospitalized cases once saturation of intensive care units is reached as $\theta^{(sat)} = 0.4 \times 1/11 = 0.0364$ (1/day). Note that consistently one obtains $\gamma_{ss}^{(sat)} = 0.6 \times 1/11 = 0.0546$ (1/day). It is assumed that there exist eight intensive care beds per 100'000 persons [6] and that 40% of the hospitalized cases need such treatment. Accordingly, saturation is reached once the number of hospitalized cases, i.e., $n_{ss}$, exceeds $C^{(\text{sat})} = 0.02\%$ of the total population.

   The adjusted rate

$$\theta(n_{ss}^{tot}) = \frac{n_s^0}{n_{ss}^{tot}} \left( \min\left\{ C^{(\text{sat})}, \frac{n_{ss}^{tot}}{n_s^0} \right\} \theta^{(0)} + \max\left\{ 0, \frac{n_{ss}^{tot}}{n_s^0} - C^{(\text{sat})} \right\} \theta^{(\text{sat})} \right) \tag{20}$$

   then quantifies the death rate of hospitalized cases as the weighted average of $\theta^{(0)}$ and $\theta^{(\text{sat})}$; the consistently adjusted recovery rate becomes

$$\gamma_{ss}(n_{ss}^{tot}) = \gamma_{ss}^{(0)} + \theta^{(0)} - \theta(n_{ss}^{tot}). \tag{21}$$

All estimates here are summarized in Table S2; note that these values can easily be adapted, if more reliable data becomes available. The resulting parameter values for our base case are provided in Table S3.

Figure 2A shows the model results without mitigation for a period of 600 days with $\mathcal{R}_0 = 2.4$. Results with $\mathcal{R}_0 \in \{1.9, 2.9\}$ are shown in Fig. S2A,B. Dashed lines represent the immune plus deceased plus infected ($n_s^0 - n_s$), dash-dotted lines the infected ($n_i^{undet} + n_i^{det}$) and solid lines the deceased ($n_d^{tot}$) population. For bigger values of $\mathcal{R}_0$, a larger immune population is needed to achieve herd immunity (right half of the graphs), and the peak in the number of infections is higher and sharper.

The plot in Fig. S2C shows the case with $\mathcal{R}_0 = 2.4$ without intensive care unit limitation; compare with Fig. 2A, which shows the same case with intensive care unit limitation (our base case). In both cases 87% of the population will become immune, which compares well with 81% infected people predicted by [6] for the UK and US populations in the absence of mitigation plans. Without intensive care the chance of dying is roughly twice as high for strong symptomatic people than with proper treatment (4.6% vs. 2.3%). While these numbers are subject to errors (mainly due to

| probability | conditional on | expression | base case value |
|---|---|---|---|
| → pre- and mild sympt. (no self isol.) | asympt. | $S^{(m)}$ | 2/3, [1/2, 2/3] |
| → strong sympt. (hospit.) | mild sympt. (self isol.) | $S^{(s)}$ | 1/5 |
| → deceased | hospit.; with icu | $M^{(0)}$ | 1/5 |
| → deceased | hospit.; no icu | $M^{(sat)}$ | 2/5 |
| **char. time scale (days)** | | | |
| → pre- and mild sympt. (no self isol.) | exposed | $S^{(m)}\beta_s^{-1}$ | 4.27 |
| → mild sympt. (self isol.) | pre- and mild sympt. (no self isol.) | $t_{im} = \xi_{ms}^{-1}$ | 1.5 |
| → strong sympt. (hospit.) | mild sympt. (self isol.) | $t_{ms} = S^{(s)}\xi_{ss}^{-1}$ | 10 |
| → deceased | str. sympt. | $M^{(0,sat)}/\theta^{(0,sat)}$ | 11 |
| → recovered | asymptomatic | $t_{ia} = \gamma_a^{-1}$ | 11.5 |
| **further parameters** | | | |
| basic reproduction number | | $\mathcal{R}_0$ | 2.4 |
| infection rate reduction factor | mild sympt. (self isol.) | $\epsilon$ | 0.1 |
| rel. intensive care capacity | | $C^{(sat)}/2.5$ | 0.00008 |
| **social distancing, contact tracing and testing** | | | |
| infection rate reduction factor | social distancing | $\lambda$ | 0 |
| testing frequency (1/days) | testing | $N^{-1}$ | 0 |
| testing process time (days) | testing | $\tau_{proc}$ | 1 |
| fraction of false negative test results | testing | $\eta$ | 0.05, $\{0.5, 0.15, 0.25\}$ |
| success rate of contact tracing | contact tracing | $\zeta$ | [0.3, 1] |
| fraction of exposed people who develop no symptoms | contact tracing | $r_1$ | [1/3, 1/2] |
| fraction of exposed people who develop no symptoms | contact tracing | $r_2$ | [0.1, 0.5] |

**Table S 2.** Estimations made for the model closure, social distancing, contact tracing and testing.

| parameter | value |
|---|---|
| $\alpha$ | 0.670 (1/day) |
| $\epsilon$ | 0.1 |
| $\beta_a$ | 0.078 (1/day) |
| $\beta_s$ | 0.156 (1/day) |
| $\gamma_a$ | 0.087 (1/day) |
| $\xi_{ms}$ | 0.667 (1/day) |
| $\gamma_{ms}$ | 0.08 (1/day) |
| $\xi_{ss}$ | 0.02 (1/day) |
| $\gamma_{ss}^{(0)}$ | 0.072 (1/day) |
| $\theta^{(0)}$ | 0.0182 (1/day) |
| $\theta^{(sat)}$ | 0.0364 (1/day) |
| **initial condition** | **value** |
| $n_s^0$ | 6'384'631'490 (world population outside of China) |
| $n_e(0)$ | 1'000 |

**Table S 3.** List of estimated parameters and initial values. Note that our model allows to easily replace any of these parameters by more precise estimates, as more data become available. The initial values of all numbers except $n_e$ are set to zero.

uncertainties in the parameter values and efforts to increase intensive care and respirator availability), it can be expected that the relevant dynamics is captured to a high degree. If the results are regarded with respect to the base case, much insight can be gained, e.g. how social distancing, mass testing and smart testing can be combined most effectively.

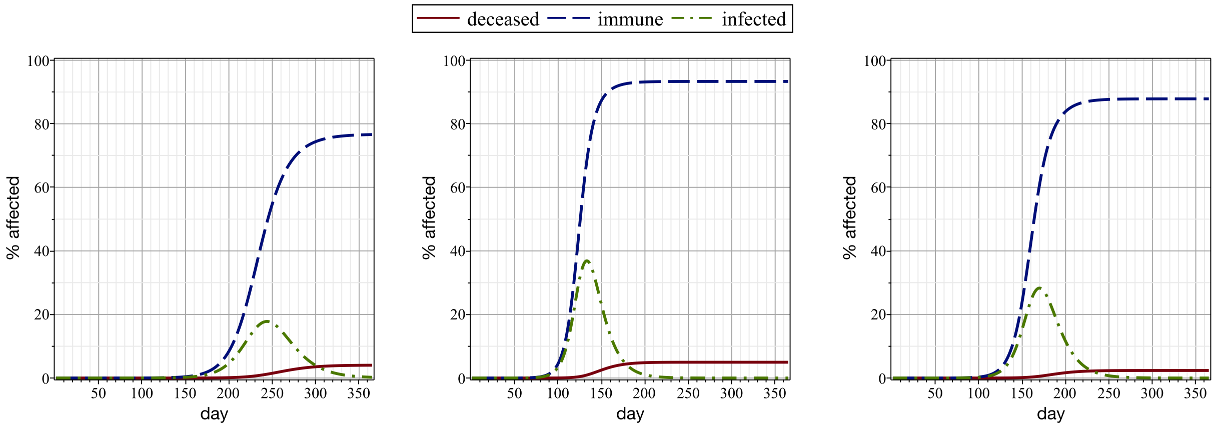


$\mathcal{R}_0$ = 1.9; with ICU capacity limit (A)

$\mathcal{R}_0$ = 2.9; with ICU capacity limit (B)

$\mathcal{R}_0$ = 2.4; without ICU capacity limit (C)

**Fig. S 2.** Alternative model outcomes when changing $\mathcal{R}_0$ of the pandemic, or relaxing the assumption that ICU beds are limiting. Dashed blue lines represent recovered and deceased, dashed-dotted green lines infected, and solid red lines deceased people. Changing $\mathcal{R}_0$ to (A) 1.9 and (B) 2.9 changes the outcomes quantitatively, but does not change the overall picture. (C) Model outcome if the assumption that full ICUs increase the death rate is dropped.

### 3. Mass Testing - How the Number of Tests Relates to $\mathcal{R}_{\text{eff}}$

Here we analyze how many tests are needed to mitigate the Covid-19 pandemic if no other mitigation strategies were applied. If we can use more tests than that, the effective reproduction number $\mathcal{R}_{\text{eff}}$ will drop below one, the incidence would decline and the pandemic would eventually end, even if no vaccines or infection therapies become available. Therefore we studied how $\mathcal{R}_{\text{eff}}$ varies once confirmed cases get isolated (in addition to self-quarantined and hospitalized individuals). Of particular value is the relationship between $\mathcal{R}_{\text{eff}}$ and the interval of testing the susceptible population (i.e., the frequency of testing needed for reducing $\mathcal{R}_{\text{eff}}$ below one). Thereby we can determine the key technical parameter of interest, i.e., the number of tests per 100'000 people that must be tested per day in order to achieve the desired $\mathcal{R}_{\text{eff}}$ value; we chose $\mathcal{R}_{\text{eff}} = 1$ as the target value for our analyses (if not indicated otherwise), which would suffice to keep the number of infected people constant.

To be realistic, we suppose that the processing time $\tau_{proc}$ of mass testing would be somewhere between half a day and two days. Furthermore, a fraction $\eta = 0.05$ of false negative test results is taken into account [19]. It should be noted, however, that this is a rough estimate. The use of standards allows for very high reproducibly of the virus RNA detection results even between different laboratories [3]. The true rate of false negatives and false positives is currently not known. We assume that a false negative rate of 5% is a conservative estimate. The current virus RNA testing capacities in continental Europe reach up to 230 tests per 100'000 people per day (e.g. in Switzerland). If equipment and supplies are not limiting for testing, e.g. by using a quantitative polymerase chain reaction (qPCR) method [1], we estimate that up to around 1'000 samples within a time frame of eight hours can be analyzed per machine. Mass testing (i.e., if $> 500 - 1'000$ tests per 100'000 perople per day would be required) could be realizable by taking advantage of next-generation RNA extraction, reverse transcription and sequencing (combined with reverse transcription and PCR)

[1] `https://www.roche.com/media/releases/med-cor-2020-03-13.htm`

to detect the virus RNA of infected people. For example, in [13] a massively parallel diagnostic assay is described for testing up to 19'200 patient samples per work flow. In principle, such very high throughput approaches can be parallelized (and potentially optimized) to provide millions of tests per day. In reality, the logistics of collecting these millions of samples would however be a major hurdle. First, we want to assess how many tests were indeed necessary to stop the virus spread (i.e., to reach $\mathcal{R}_{\text{eff}} \leq 1$ if no other mitigation strategies were applied).

Obviously, the scenario that the whole susceptible population is tested perfectly at once would lead to a trivial disease free state. However, this is an unrealistic scenario, not only because of a lack of test capacity, but also due to logistic and compliance concerns. Therefore, it is only realistic to assume that individuals would be tested at different schedules. Let us consider a situation where each person is tested once every $N$ days. Note that this is equivalent to testing a random fraction of $1/N$ of the susceptible population every day. Therefore we focus on the set $\{1, ..., N\}$ of days. An individual is infected at some random time $x$. To characterize $x$, we assume that the likelihood of getting infected does not vary much during these days, which is justified if testing is applied at an intensity such that $\mathcal{R}_{\text{eff}} \approx 1$. Hence $x$ becomes uniformly distributed in the interval $[1, N]$. The time delay between infection and detection would then be $\tau_{det} = N - x + \tau_{proc}$. Finally, we sample the latency time $x_l$ from a log-normal distribution with $5.34 \pm 2.7249$ (see Section 2).

Intuitively, by conducting mass testing on individuals who are neither self-quarantined nor hospitalized, positive cases will be detected from exposed, asymptomatic and mild-symptomatic compartments. To quantify each detection rate, it is essential to compare detection time versus the latency period; therefore we consider the effect of testing on these three compartments individually:

1. *Exposed*: Once testing occurs during the latency period of an infected individual, they would be detected from the exposed compartment. This translates into an event set $\mathcal{A} : \tau_{det} \leq x_l$. The rate of detecting individuals by testing from the exposed population then reads

$$k_e = (1 - \eta)\mathcal{P}_{\mathcal{A}}\mathbb{E}_{\mathcal{A}}\left[\tau_{det}^{-1}\right], \quad (22)$$

where $P_{\mathcal{A}}$ and $\mathbb{E}_{\mathcal{A}}$ denote frequency of such events and conditional expectation, respectively.

2. *Mild-symptomatic*: Once testing occurs after the latency period, one has to distinguish between two types of infection developments. According to our setting, two thirds of the infected individuals would develop symptoms that will lead them to self-isolate. Please note, that the fraction of infected individuals that remain asymptomatic may range between 33%-50% [6, 7, 27]; see Table S2. Therefore, we have also tested scenarios where 60% or 50% of infected will will progress to develop symptoms; see Fig. S5. These individuals may in fact turn to testing centers in order to detect the virus and incentivize the decision for self-quarantine. They can be detected by testing and therefore sent into quarantine in the span of one and a half days after becoming infectious. The relevant event set is $\mathcal{B} : (\tau_{det} \geq x_l) \cap (\tau_{det} \leq (x_l + 3/2))$, from which one obtains

$$k_s = \frac{2}{3}(1 - \eta)\mathcal{P}_{\mathcal{B}}\mathbb{E}_{\mathcal{B}}\left[\tau_{det}^{-1}\right] \quad (23)$$

for the test detection rate of mild-symptomatic persons.

3. *Asymptomatic*: This is arguably the most important group to consider, because they will not know that they are infected and they can make up as much as 50% of the entire group of the infected people. Importantly, they are very hard to identify using the current mitigation strategies, including app based contact tracing [7]. We

will discuss this in more detail, below. Testing may catch asymptomatic cases. These individuals won't have symptoms and would then recover after 11.5 days. In their first one and a half days they share same the time-line as mild symptomatic ones. Therefore, in this scenario two event sets $\mathcal{B} : (\tau_{det} \geq x_l) \cap (\tau_{det} \leq (x_l + 3/2))$ and $C : ((\tau_{det} \geq (x_l + 3/2)) \cap (\tau_{det} \leq (x_l + 11.5))$ become relevant. We obtain the detection rate from the asymptomatic compartment as

$$k_a = \frac{1}{3}(1-\eta)\mathcal{P}_{\mathcal{B}}\mathbb{E}_{\mathcal{B}}\left[\tau_{det}^{-1}\right] + (1-\eta)\mathcal{P}_C\mathbb{E}_C\left[\tau_{det}^{-1}\right]. \tag{24}$$

Before finding the map from testing frequency $N^{-1}$ to $\mathcal{R}_{\text{eff}}$, we need to find out how $\mathcal{R}_{\text{eff}}$ varies with respect to the test detection rates $k_e$, $k_a$ and $k_s$. Hence, let us define $\mathcal{R}_{\text{eff}}^{wt}$ as the reproduction number subject to testing. To compute $\mathcal{R}_{\text{eff}}^{wt}$, the main dynamics of the infected population, which can be described by $[n_e(t), n_{ia}(t), n_{im}(t), n_{ms}(t), n_{ss}(t), \tilde{n}_e(t), \tilde{n}_{ia}(t), \tilde{n}_{im}(t), \tilde{n}_{ms}(t), \tilde{n}_{ss}(t)]^T$, is split into the rate of appearance $f$ of new infected individuals and transfer $\tilde{V}$ of infected ones across different compartments, i.e.,

$$f = \begin{bmatrix} 0 & \alpha/2 & \alpha & \epsilon & 0 & 0 & 0 & 0 & 0 & 0 \\ 0 & 0 & 0 & 0 & 0 & 0 & 0 & 0 & 0 & 0 \\ 0 & 0 & 0 & 0 & 0 & 0 & 0 & 0 & 0 & 0 \\ 0 & 0 & 0 & 0 & 0 & 0 & 0 & 0 & 0 & 0 \\ 0 & 0 & 0 & 0 & 0 & 0 & 0 & 0 & 0 & 0 \\ 0 & 0 & 0 & 0 & 0 & 0 & 0 & 0 & 0 & 0 \\ 0 & 0 & 0 & 0 & 0 & 0 & 0 & 0 & 0 & 0 \\ 0 & 0 & 0 & 0 & 0 & 0 & 0 & 0 & 0 & 0 \\ 0 & 0 & 0 & 0 & 0 & 0 & 0 & 0 & 0 & 0 \\ 0 & 0 & 0 & 0 & 0 & 0 & 0 & 0 & 0 & 0 \end{bmatrix} \tag{25}$$

and

$$\tilde{V} = \begin{bmatrix} \beta_a + \beta_s + k_e & 0 & 0 & 0 & 0 & 0 & 0 & 0 & 0 & 0 \\ -\beta_a & \gamma_a + k_a & 0 & 0 & 0 & 0 & 0 & 0 & 0 & 0 \\ -\beta_s & 0 & \xi_{ms} + k_s & 0 & 0 & 0 & 0 & 0 & 0 & 0 \\ 0 & 0 & -\xi_{ms} & \xi_{ss} + \gamma_{ms} & 0 & 0 & 0 & 0 & 0 & 0 \\ 0 & 0 & 0 & -\xi_{ss} & \theta + \gamma_{ss} & 0 & 0 & 0 & 0 & 0 \\ -k_e & 0 & 0 & 0 & 0 & \beta_a + \beta_s & 0 & 0 & 0 & 0 \\ 0 & -k_a & 0 & 0 & 0 & -\beta_a & \gamma_a & 0 & 0 & 0 \\ 0 & 0 & -k_s & 0 & 0 & -\beta_s & 0 & \xi_{ms} & 0 & 0 \\ 0 & 0 & 0 & 0 & 0 & 0 & 0 & -\xi_{ms} & \xi_{ss} + \gamma_{ss} & 0 \\ 0 & 0 & 0 & 0 & 0 & 0 & 0 & 0 & -\xi_{ss} & \theta + \gamma_{ss} \end{bmatrix}, \tag{26}$$

which leads to

$$\mathcal{R}_{\text{eff}}^{wt} = \rho(f\tilde{V}^{-1}) = \frac{\alpha\beta_s}{\beta_a + \beta_s + k_e}\left[\frac{\beta_a}{2(\gamma_a + k_a)\beta_s} + \frac{1}{\xi_{ms} + k_s}\left(1 + \frac{\epsilon\xi_{ms}}{\gamma_{ms} + \xi_{ss}}\right)\right]. \tag{27}$$

It is important to emphasize that in contrast to social distancing the effect of testing on the reproduction number is not through reducing the infection rate (i.e., $f$ remains the same), but rather through transfer of infected individuals to quarantine, either at home or in hotel rooms, or in care units with strict hygiene barriers, all of which would reduce the likelihood of transmission (i.e., changing $V$ to $\tilde{V}$).

Using Monte-Carlo to estimate the test detection rates given by Eqs. (22) and (23) one can compute the ratio $\mathcal{R}_{\text{eff}}^{wt}/\mathcal{R}_0$ with respect to the testing frequency $N^{-1}$. The plots in Figs. S3A,B show the number of tests required to reach $\mathcal{R}_{\text{eff}} = 1$. It depends on the time from sampling to result (note: we assume immediate notification and immediate implementation of quarantine measures upon notification) and on the false negative rate (5% in Fig. S3A and 15% in Fig. S3B). The horizontal green dashed lines indicate $\mathcal{R}_{\text{eff}}^{wt} = 1$, if the virus reproduction rate in the case without any mitigation is $\mathcal{R}_0 = 2.4$.

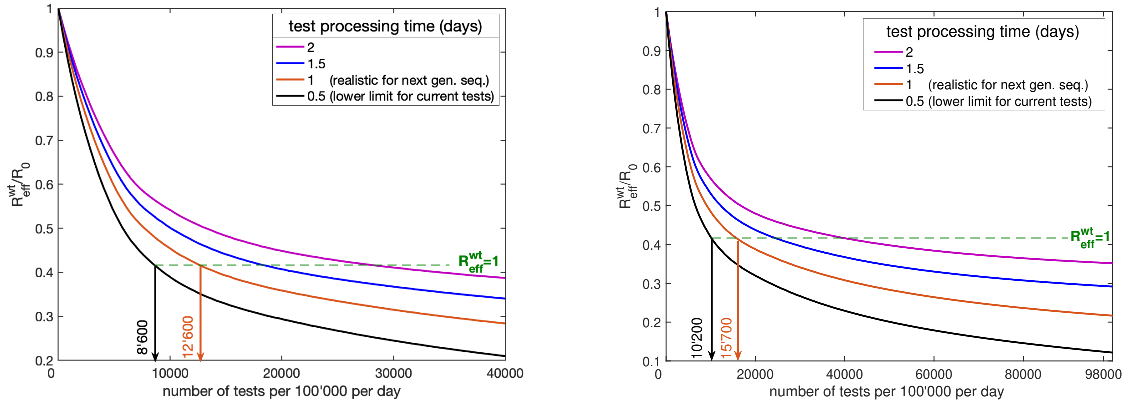


(A) (B)

**Fig. S 3.** Mass testing: The number of tests required to reach $\mathcal{R}_{\text{eff}} = 1$ depends on the time from sampling to result. A mitigation strategy relying on mass testing alone is assumed and we computed the number of tests performed per day, which are needed to achieve a particular test-speed dependent $\mathcal{R}_{\text{eff}}^{wt}/\mathcal{R}_0$ ratio; for (A) 5% and (B) 15% false negative test results are assumed. The green lines indicate $\mathcal{R}_{\text{eff}}^{wt} = 1$. Test speeds were: 0.5 days (black line), 1 day (orange line), 1.5 days (blue line) and 2 days (purple line).

One can see for example that reducing the initial $\mathcal{R}_0 = 2.4$ to one would require to test the entire susceptible population roughly once every eight days, if the false negative rate is 5%, if testing results are available after one day and quarantine of the detected individuals commences immediately. Corresponding results are depicted in Fig. S4B, which shows model outcomes if $\mathcal{R}_{\text{eff}}$ is reduced to one by testing 12'600 per 100'000 people per day. Note that this corresponds to a testing interval of 7.92 days or equivalently to a fraction of 1/7.92 which has to be tested every day. Figure S4A shows the outcome when social distancing is applied to reduce $\mathcal{R}_{\text{eff}}$ to one, which is equivalent to a 58% reduction of the infection rate. We define this as moderate social distancing; see Table S1. By comparing Figs. S4A and S4B one observes that mass testing and social distancing yield qualitatively equivalent results. For half a day delay time the testing interval can be increased to roughly twelve days and for a delay time of one and a half days it would be five days. This information is important for optimizing technical development decisions. The overall testing capacity needs to be larger, if the testing method requires more time (Fig. 3) or if delays in case-notification or implementation of quarantine measures occur. This could be justified, if a low-cost technique (such as next generation sequencing) could be devised [13]. Alternatively, one could aim for fewer tests, if they are completed in less than half a day. If combined with social distancing, the effective reproduction number can be reduced by the factor $(1 - \lambda)\mathcal{R}_{\text{eff}}^{wt}/\mathcal{R}_0$, where $\lambda \in [0, 1]$ is the intensity of social distancing ($\lambda = 0$ means no social distancing and $\lambda = 1$ means complete isolation of everybody).

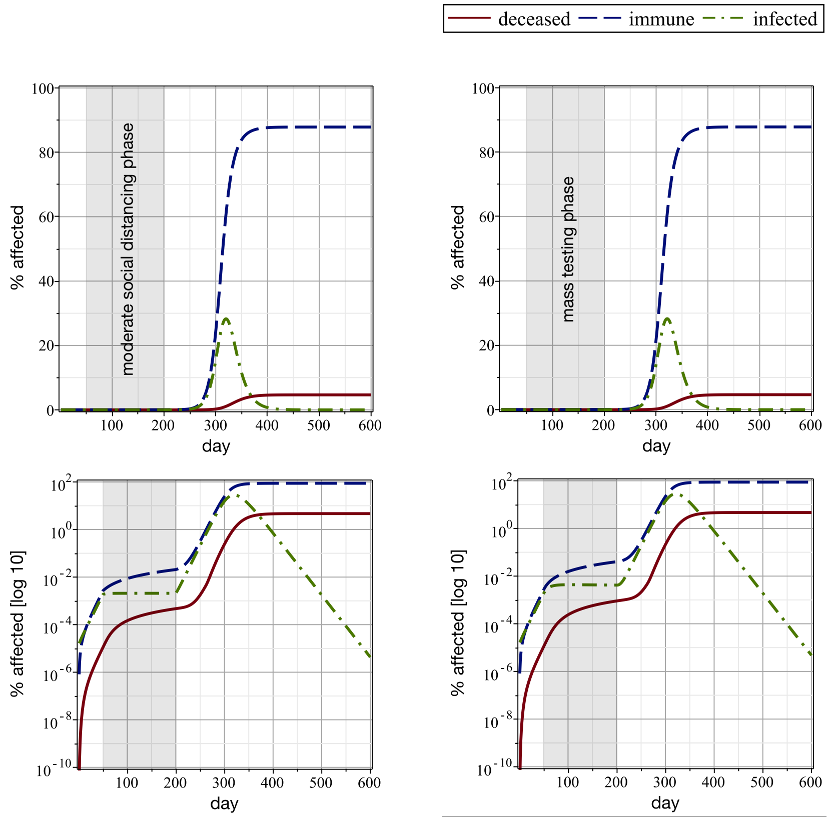


moderate social distancing
($\mathcal{R}_{\text{eff}} = 1$)
(A)

mass testing 12'600 per 100'000 per day
($\mathcal{R}_{\text{eff}} = 1$)
(B)

**Fig. S 4.** Moderate social distancing vs. mass testing: Model outcomes if $\mathcal{R}_{\text{eff}}$ is reduced to 1 by mass testing or social distancing. (A) Moderate social distancing ($\mathcal{R}_{\text{eff}} = 1$, equivalent to a 58% reduction in infection rate) and (B) testing 12'600 per 100'000 people per day yield qualitatively equivalent results.

### 4. Contact Tracing - How Size and Infectiousness of the Asymptomatic Population Relates to $\mathcal{R}_{\text{eff}}$

Contact tracing has been proposed to slow down or even stabilize the pandemic [7, 12, 8]. The strategy is that symptomatic individuals who go into self quarantine will use an App to alert all proximity contacts of the past two weeks. Subsequently, these identified individuals self-isolate themselves. For the following analysis we assume that the notice-to-quarantine time delay is negligible [7, 12].

To analyze the effect of contact tracing on the effective reproduction number, the expected infectiousness $A(\tau)$ at time $\tau$ after infection plays a central role. By knowing $A(\tau)$ we can extract the basic reproduction number as

$$\mathcal{R}_0 = \int_0^\infty A(\tau) d\tau. \tag{28}$$

Note that the above equation is consistent with our previous computation of $\mathcal{R}_0$ in the linear regime [4]. In our compartmental setting shown in Fig. 1 one obtains the expression

$$A(\tau) = \sum_{i \in \mathcal{S}_c} \mathcal{P}_i A_i(\tau), \tag{29}$$

where $\mathcal{P}_i$ is the probability that an infected individual is in compartment $c_i$, $\mathcal{S}_c = \{s, e, ia, ra, im, ms, rs, ss, d\}$ is the index set of all compartments and $A_i(\tau)$ denotes the infectiousness at time $\tau$ after infection, if the individual is in $c_i$. Correspondingly, we denote the time spent in each compartment as $t_{i \in \mathcal{S}_c}$. From Eqs. (28) and (29), if we assume that infectiousness is constant inside each compartment, we obtain

$$\mathcal{R}_0 = \overbrace{\alpha r_1 r_2 t_{ia}}^{\mathcal{R}_0^{asym}} + \overbrace{\alpha (1 - r_1) \left( t_{im} + \frac{1}{10} t_{ms} \right)}^{\mathcal{R}_0^{sym}}, \tag{30}$$

where $r_1 = 1 - S^{(m)}$ is the fraction of exposed people who develop no symptoms (thus remain asymptomatic), and $r_2$ is the factor by which asymptomatic people are less infectious than symptomatic ones. For both $r_1$ and $r_2$ different values are suggested in the literature. For $r_1$ one finds 1/3 in [6], 0.4 in [7] and 0.5 in [27], and for $r_2$ one finds 0.1 in [7], 2/3 in [6] and 1 in [14]. Based on these published numbers we consider $r_1 \in [0.3, 0.5]$ and $r_2 \in [0.1, 0.5]$. From Eq. (30) with our base case values $r_1 = 1/3$ and $r_2 = 0.5$ one obtains $\alpha = 0.67$ (1/day), which is consistent with our previous parameter estimation. Based on our previous assumptions and on the values in Table S2 we obtain $t_{ia} = \gamma_a^{-1} = 11.5$ days, $t_{im} = \xi_{ms}^{-1} = 1.5$ days and $t_{ms} = S^{(s)} \xi_{ss}^{-1} = 10$. Interesting here is the fraction

$$\kappa = \frac{\mathcal{R}_0^{sym}}{\mathcal{R}_0} = \frac{(1 - r_1)(t_{im} + t_{ms}/10)}{r_1 r_2 t_{ia} + (1 - r_1)(t_{im} + t_{ms}/10)} \tag{31}$$

of infected people who got infected by symptomatic cases, since this is the maximum relative reduction of $\mathcal{R}_0$ which can be achieved by tracing contacts of symptomatic individuals (by classical contact tracing or by using an app) with a success rate of $\zeta = 1$. Ignoring secondary infections (their probability becomes around 0.8%) we obtain the approximation

$$\mathcal{R}_{\text{eff}}^{ct} \approx [1 - \zeta \kappa] \mathcal{R}_0 \tag{32}$$

for the effective reproduction number, if contact tracing is employed. Figures S5A and S5B show the performance of contact tracing for different $r_1$-, $r_2$- and $\zeta$-values. The numbers attached to the isolines refer to the ratio $\mathcal{R}_{\text{eff}}^{wt}/\mathcal{R}_0$,

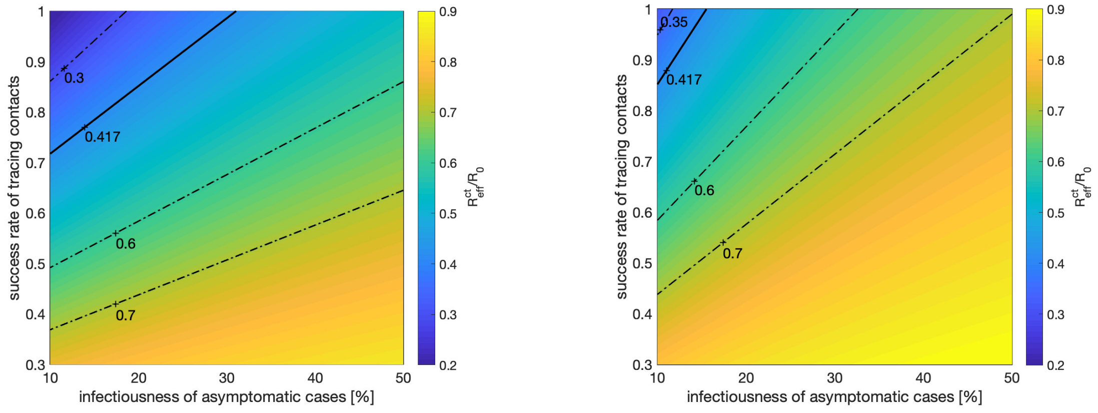


(A) (B)

**Fig. S 5.** Contact tracing: Effectiveness of contact tracing as function of relative infectiousness of asymptomatic cases ($r_2$) and the success rate of contact tracing ($\zeta$). (A): 33% of infected ones are asymptomatic ($r_1 = 0.33$); (B): 50% of infected ones are asymptomatic ($r_1 = 0.5$). The numbers attached to the isolines refer to the ratio $\mathcal{R}^{wt}_{\text{eff}}/\mathcal{R}_0$, and the bold contours depict combinations of $r_2$ and $\zeta$ for which $\mathcal{R}_{\text{eff}}$ is reduced from 2.4 to $\mathcal{R}^{ct}_{\text{eff}} = 1$. In all computations, notice-to-quarantine time was neglected and it was assumed that quarantined contacts are not infectious.

and the bold contours depict combinations of $r_2$ and $\zeta$ for which $\mathcal{R}_{\text{eff}}$ is reduced from 2.4 to $\mathcal{R}^{ct}_{\text{eff}} = 1$. One can conclude that the effect of contact tracing, if only applied to identify contacts with symptomatic persons, is limited to optimistic assumptions concerning the parameters dictating the Covid-19 pandemic and strongly depends on size and infectiousness of the asymptomatic population relative to size and infectiousness of the symptomatic one. Even if the most optimistic assumptions would hold, more substantial reductions of $\mathcal{R}_{\text{eff}}$ would be desirable in order to accelerate the end of the pandemic (e.g. even in the absence of effective therapies or vaccines). For our base parameters of $r_1 = 1/3$ and $r_2 = 1/2$, contact tracing alone would not lead to an effective reproduction number of one (see Fig. S 5A at 50% infectiousness).

### 5. Smart Testing - How Selectivity Relates to $\mathcal{R}_{\text{eff}}$

Here it is studied how the number of required tests can be reduced, if one is able to identify (and propose testing to) a subpopulation with a prevalence higher than the overall population.

Without any additional knowledge, to achieve the discussed detection rates $k_e$, $k_a$ and $k_s$ and corresponding reductions in $\mathcal{R}_{\text{eff}}$, one has to test the entire undetected population once every $N$ days (or equivalently every day a random fraction of $1/N$). To improve the efficiency of testing, i.e., the probability per test of getting a positive result by avoiding unnecessary testing of people who have a low likelihood of being infected, one can reduce the sample population by the following approaches:

1. *Serological testing*: With serological testing one can remove the recovered population from the pool of undetected individuals, and thus the same number of positive test results can be achieved with fewer tests. However, during an early stage of the pandemic the relative size of the undetected recovered compared to the whole undetected population is very small, and therefore the gain would be negligible. Nevertheless, this approach can easily be integrated into any mass testing strategy once reliable serological tests become available. Here, we can assume

that immune individuals will remain immune for an extended period of time (e.g. up to 1-2 years; however, this is still subject to verification). If immune people cannot be infected for a second time (or are infected at a much lower rate than susceptible individuals), one can collect the information on positive test results to exclude the immune individuals from the Covid-19 RNA testing.

2. *Inference from contact tracing*: A more effective approach would be based on an inference model (e.g. by using contact tracing of infected individuals), which allows to divide the sample population $\mathcal{D}$ into one subpopulation $\tilde{\mathcal{D}}$ with a higher and the remainder with a lower percentage of infected individuals. For the following analysis we denote the size of $\mathcal{D}$ with $n$ and that of $\tilde{\mathcal{D}}$ with $\tilde{n}$. Further, $\tilde{p}$ is the fraction of infected persons in $\tilde{\mathcal{D}}$ and $p$ that in $\mathcal{D}$. Testing every person in $\tilde{\mathcal{D}}$ at a frequency of $1/N$ would require $\tilde{n}/N$ tests per day; opposed to $n/N$ tests, if the whole sample population was tested. The respective numbers of positive test results per day, on the other hand, would be $\tilde{P}(N) = \tilde{p}\tilde{n}/N$ opposed to $P(N) = pn/N$. In order to obtain the same number of positive results from the subpopulation $\tilde{\mathcal{D}}$ as one would get from $\mathcal{D}$, one has to reduce the test interval $N$ to $\tilde{N}$, such that $\tilde{P}(\tilde{N}) = P(N)$. From this one obtains $\tilde{N} = N(\tilde{p}\tilde{n})/(pn)$ and one can conclude that the number of tests required to achieve the same overall quota reduces by the factor

$$r = \frac{\tilde{n}/\tilde{N}}{n/N} = \frac{N\tilde{n}}{\tilde{N}n} = \frac{p}{\tilde{p}}. \tag{33}$$

In order for this result to be practically meaningful, $\tilde{N}$ has to be at least one, which translates into the requirement that

$$\frac{\tilde{n}}{n} \geq \frac{r}{N}. \tag{34}$$

In short, if one can identify a subpopulation $\tilde{\mathcal{D}} \subset \mathcal{D}$ for which the percentage of infections is higher by a factor of $r^{-1}$ than in $\mathcal{D}$, and which is larger than $nr/N$, then the number of tests needed to obtain the same reproduction number reduces by the factor $r$. The curves in Fig. 3B show the relationship between number of tests per 100'000 people per day needed to achieve $\mathcal{R}_{\text{eff}} = 1$ and the prevalence ratio between sub- and overall population; in combination with mild social distancing (solid line) and without social distancing (dashed line).

### 6. Contact Counting - How to Screen Large Enough Subpopulations with High Prevalence

Here we devise a way to screen large enough subpopulations with a much higher prevalence and infectiousness than the overall population, which is a prerequisite for smart testing.

We study three subpopulations as potential candidates for our smart testing mitigation approach; of interest are their prevalence, their infectiousness and their size. Next we describe them and provide quantitative estimates for the most relevant subpopulation.

1. *Contacts of symptomatic cases:* A straight-forward approach would be to choose the contacts of symptomatic cases as our subpopulation. While this group is highly likely to be infected, this approach has one major drawback. In fact, the outcome of testing contacts of symptomatic cases would not be much different than that of contact tracing mentioned before (and discussed in detail by Ferretti et al.[7]). Therefore, it suffers from the same limitation of not catching sufficient numbers of asymptomatic infections. Besides tracing contacts which potentially got infected by a symptomatic individual, one may also find the contact by whom it got infected. That person has most likely recovered, since he/she got infected roughly 10-14 days ago. Therefore this contact would not be tested positive (as virus titers may already be low and as we still lack reliable serological tests) and hence testing contacts of symptomatic ones would not lead us to a larger group with a sufficient number of asymptomatic cases.

2. *Direct and indirect contacts of symptomatic cases:* One way to cope with the issue arising from lack of enough asymptomatic cases in the contacts of symptomatic ones is to enlarge our sample population and include also indirect contacts of symptomatic individuals in the past two weeks. This strategy, while most probably catching enough asymptomatic cases, may not reduce the burden of mass testing, since now the size of the subpopulation becomes simply too large. This problem has also been noted by others [12]. For example at a prevalence of 1% in the total population, and assuming 10 contacts per person, the size of this subpopulation becomes almost as large as the whole population.
3. *High-contact individuals* (Fig. 4A): In this scenario we only test those with significantly more contacts than the average. In the following we show that indeed this strategy allows to screen a high prevalence subpopulation which is also large enough to stop the pandemic. It is also important to emphasize that the prerequisite of this strategy is to utilize a contact counter, which may be integrated into an existing contact tracing app that uses bluetooth technology.

While the improvements resulting from contact counting can be estimated based on the prevalence ratio, the contact counting scheme has a more fundamental feature that exhibits itself directly in the effective reproduction number. In fact by cutting out the highly transmissive parts of the population network, we reduce the effective reproduction number significantly. This reduction in $\mathcal{R}_{\text{eff}}$ can be evaluated by considering the transmissibility of the disease $T$. In short, considering a normalized recovery rate, $T$ is the probability that an infected person infects one of their contacts per unit of time. Consider $\mathcal{K}$ to be the degree of connectivity of a person; therefore we can compute $T$ for a heterogeneous network as [21, 24, 25]

$$T = \mathcal{R}_0 \frac{\mathbb{E}[\mathcal{K}]}{\mathbb{E}[\mathcal{K}^2 - \mathcal{K}]}. \tag{35}$$

Now imagine a scenario where we halt the virus-spread among all individuals with a degree of connectivity above $\mathcal{K}_0$; for example via vaccinating every person who has contact numbers above $\mathcal{K}_0$. Therefore, we get a reduction in the virus reproduction number

$$\mathcal{R}_{\text{eff}}^{nn} = \frac{\mathbb{E}[\mathcal{K}^2 - \mathcal{K}|\mathcal{K} \leq \mathcal{K}_0]}{\mathbb{E}[\mathcal{K}^2 - \mathcal{K}]} \frac{\mathbb{E}[\mathcal{K}]}{\mathbb{E}[\mathcal{K}|\mathcal{K} \leq \mathcal{K}_0]} \mathcal{R}_0, \tag{36}$$

where $\mathcal{R}_{\text{eff}}^{nn}$ denotes the effective reproduction number of the new network. For a specified size of the new network, this results in a significant reduction of the reproduction number as the tail of the contact distribution is removed (Fig. 4B). However, in practice there are failures in containing the virus-spread through highly connected people. The efficacy $\mathcal{E}$ of a smart testing based on contact counting thus depends on the number of highly connected people who would employ the contact counting app, as well as the accuracy of the tests. In the following we compute how these boundary conditions affect STeCC. However, before proceeding, notice that we suppose that the isolated individuals have negligible contributions to the virus-spread. Furthermore, we assume that elderly people (above 70 years of age) are shielded by isolation and that children below 10 years would not contribute to the infection dynamics [29, 11]. Therefore our target subpopulation is considered to be in possession of smart-phones.

Let us define a testing regime, where we screen through app users with number of connections $\mathcal{K} \geq \mathcal{K}_0^{(1)}$ at day 1, $\mathcal{K}_0^{(2)} \leq \mathcal{K}_0 \leq \mathcal{K}_0^{(1)}$ at day 2 and so-forth until testing $\mathcal{K}_0^{(T_t)} \leq \mathcal{K}_0 \leq \mathcal{K}_0^{(T_t-1)}$ at day $T_t$. Consequently, based on the test results, we ask the positively tested individuals to quarantine themselves. We fix the testing cycle to $T_t = 7$ days (see Fig. 4). Consider $\zeta$ to be the fraction of smart-phone owners who utilize the app. The portion of the network besides the fraction $\zeta$ that we can disconnect from the population depends on the probability of the event that a positively tested highly connected individual could pass on the virus to at least one person in the past $T_t + \tau_{proc}$ days.

Let us denote such an event by $\mathcal{A}_{out}$ and consider the test processing time $\tau_{proc}$. Therefore the reduction in $\mathcal{R}_{\text{eff}}$ during the whole cycle of STeCC becomes

$$\mathcal{R}^{nn}_{\text{eff}} = \frac{\mathcal{P}_{\mathcal{K}\leq\mathcal{K}_0^{(T_t)}}\, \mathbb{E}[\mathcal{K}^2 - \mathcal{K}|\mathcal{K} \leq \mathcal{K}_0^{(T_t)}] + (1-\mathcal{E})\mathcal{P}_{\mathcal{K}\geq\mathcal{K}_0^{(T_t)}}\, \mathbb{E}[\mathcal{K}^2 - \mathcal{K}|\mathcal{K} \geq \mathcal{K}_0^{(T_t)}]}{\mathcal{P}_{\mathcal{K}\leq\mathcal{K}_0^{(T_t)}}\, \mathbb{E}[\mathcal{K}|\mathcal{K} \leq \mathcal{K}_0^{(T_t)}] + (1-\mathcal{E})\mathcal{P}_{\mathcal{K}\geq\mathcal{K}_0^{(T_t)}}\, \mathbb{E}[\mathcal{K}|\mathcal{K} \geq \mathcal{K}_0^{(T_t)}]} \frac{\mathbb{E}[\mathcal{K}]}{\mathbb{E}[\mathcal{K}^2 - \mathcal{K}]}\mathcal{R}_0, \tag{37}$$

where $\mathcal{E}$ is the efficacy of STeCC and can be computed via

$$\mathcal{E} = 1 - ((\mathcal{P}_{\mathcal{A}_{out}}(1-\eta) + \eta)\zeta + (1-\zeta)). \tag{38}$$

Note that $\eta$ is the fraction of false negative test results , $\mathcal{P}_{\mathcal{A}_{out}}$ is the probability of the event $\mathcal{A}_{out}$, $\mathcal{P}_{\mathcal{K}\leq\mathcal{K}_0^{(T_t)}}$ is the probability that an individual has contacts below $\mathcal{K}_0^{(T_t)}$ and $\mathcal{P}_{\mathcal{K}\geq\mathcal{K}_0^{(T_t)}} = 1 - \mathcal{P}_{\mathcal{K}\leq\mathcal{K}_0^{(T_t)}}$. It is evident that $\mathcal{R}^{nn}_{\text{eff}}$ and $\mathcal{E}$ would now depend on the network topology and latency time, respectively.

#### *6.1. Network Topology*

In order to model the heterogeneity of a population relevant for disease modeling, scale-free networks offer appropriate features [5, 21, 18, 25]. Scale-free networks are characterized by a power-law distribution which determines the probability density function $P_k$ of the degree of connectivity $k$ per node. We employ a continuous approximation [26], that is,

$$P_k = \alpha_p k^{-(2+\gamma)} \quad (m \leq k \leq k_c) \;\; \& \;\; (0 \leq \gamma \leq 1), \tag{39}$$

where

$$\alpha_p = \frac{(1+\gamma)m^{1+\gamma}}{1-(k_c/m)^{-(1+\gamma)}}. \tag{40}$$

Notice that by contacts we mean disease relevant contacts. We adopt $\gamma = 0.3$ and fix the upper and lower cut-offs by $k_c = 552$ and $m = 4$, respectively. These choices were made in order to have a realistic range for number of contacts and obtain an average number of contacts of $\mu = 13.4$, which is consistent with the data provided in [23]. Two extreme choices of $\gamma$ include $\gamma = 1$ and $\gamma = 0$. While the former leads to the celebrated Barabási and Albert (BA) model [1]; the latter has been used in disease spread models, e.g. see [21]. Note that we conducted a sensitivity analysis of our results with respect to $\gamma$, see Figs. S7 and S8.

With the choice of the degree of connectivity distribution we can compute the relative size of the population with a degree of connectivity above a certain $\mathcal{K}_0^{(T_t)}$:

$$\mathcal{P}_{\mathcal{K}\geq\mathcal{K}_0^{(T_t)}} = \frac{\alpha_p}{1+\gamma}\left(\mathcal{K}_0^{(T_t)-(1+\gamma)} - k_c^{-(1+\gamma)}\right). \tag{41}$$

Before proceeding further, let us mention that for the perfect efficacy $\mathcal{E} = 1$, we can reach $\mathcal{R}^{nn}_{\text{eff}} = 1$ with $\mathcal{K}_0^{(T_t)} = 138.8$. This accounts for 0.83% of the population with highest degree of connectivity. Now, to translate $\mathcal{P}_{\mathcal{K}\geq\mathcal{K}_0^{(T_t)}}$ of the population into the equivalent number of tests, suppose we conduct $N_{test}$ per day per 100'000 people. Thus we obtain

$$\mathcal{K}_0^{(T_t)} = \left(\frac{(1+\gamma)N_{test}T_t}{10^5\alpha_p\zeta} + k_c^{-(1+\gamma)}\right)^{-1/(\gamma+1)}. \tag{42}$$

#### 6.2. *Uncontained Virus Spread*

Since we test these high-contact individuals once every $T_t = 7$ days (test cycle depicted in Fig. 4B), there is a chance that a virus-positive person has already transmitted the virus between two successive tests. The probability of such events depends on the latency time $\tau_l$ and average number of contacts of the individual. The infection time $\tau_0$ is uniformly distributed between 0 and $T_t$. Now, since these individuals have contacts way above average, we take a conservative estimation that

$$\mathcal{P}_{\mathcal{A}_{out}} = \text{Prob}\Big\{(\tau_0 + \tau_1) \leq (T_t + \tau_{proc})\Big\}, \tag{43}$$

which becomes 0.4296 for the case of $T_t = 7$ (day) and $\tau_{proc} = 1$ (day).

#### 6.3. *Scenario Analysis*

Now we are ready to consider the following scenarios and compute the reduction of the effective reproduction number as a function of number of app users.

1. *Scneario A:* In this STeCC alone scenario we identify high-contact individuals in cycles of 7 days (Fig. 4B), ask them to be tested and ask the positively tested individuals to go into self-quarantine. We compute the reduced effective reproduction number $\mathcal{R}_{\text{eff}}^{ST-A}$ using Eq. (37) with the efficacy equation (38). For our base parameters together with the network topology with $\gamma = 0.3$, $m = 4$ and $k_c = 552$, Fig. S 6 shows the performance of STeCC-A. While up to 30% reduction of $\mathcal{R}_{\text{eff}}$ can be achieved with a combination of 90% app users among smart phone users (which would only be achieved in very optimistic scenarios) and 200 tests per 100'000 per day, it is clearly insufficient to halt the pandemic. The reason lies in the fact that we would not be able to contain the virus spread from almost 40% of highly connected people resulting from the transmission events that occur between their infection and the date of the virus test. Next, we introduce a combination that can significantly reduce this virus spread.

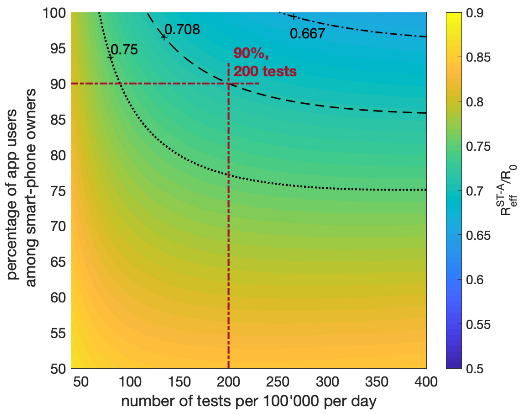


**Fig. S 6.** STeCC alone scenario (STeCC-A): $\mathcal{R}_{\text{eff}}^{ST-A}/\mathcal{R}_0$ as function of number of smart tests per 100′000 per day and the percentage of smart-phone owners who participate in STeCC (Scenario A). For $\mathcal{R}_0 = 2.4$, the dotted line indicates $\mathcal{R}_{\text{eff}}^{ST-A} = 1.8$, the dashed line $\mathcal{R}_{\text{eff}}^{ST-A} = 1.7$ and the dashed-dotted line $\mathcal{R}_{\text{eff}}^{ST-A} = 1.6$.

2. *Scenario B:* Here we consider a variant of STeCC in which besides the positively tested ones also their contacts are asked to quarantine. This results in a much lower effective reproduction number of $\mathcal{R}_{\text{eff}}^{ST-B}$, which is obtained from Eq. (37) with the improved efficacy

$$\mathcal{E} = \mathcal{E}^{ST-B} = 1 - \left(\left(\mathcal{P}_{\mathcal{A}_{out}}(1-\eta)\overbrace{(1-\zeta)}^{\text{due to quarantine}} + \eta\right)\zeta + (1-\zeta)\right). \tag{44}$$

The results shown in Fig. 4C predict a much stronger effect than STeCC alone (Fig. S 6). The combination of 90% user percentage and almost 400 tests per 100'000 per day now lead to $\mathcal{R}_{\text{eff}}^{ST-B} = 1$. Again, this approach alone will likely not suffice to halt the pandemic under realistic conditions.

3. *Scenario C:* Since the app already provides the contacts, we can further improve the STeCC based mitigation by combining it with conventional contact tracing (see §4). This is especially interesting since neither contact tracing nor STeCC alone are effective enough to halt the pandemic under realistic conditions. Therefore, on top of scenario B, we also ask the contacts of symptomatic cases to quarantine, which leads to an effective reproduction number of

$$\mathcal{R}_{\text{eff}}^{ST-C} = \frac{\mathcal{P}_{\mathcal{K}\le\mathcal{K}_0^{(T_t)}}\,\mathbb{E}[\mathcal{K}^2-\mathcal{K}|\mathcal{K}\le\mathcal{K}_0^{(T_t)}] + (1-\mathcal{E}^{ST-B})\mathcal{P}_{\mathcal{K}\ge\mathcal{K}_0^{(T_t)}}\,\mathbb{E}[\mathcal{K}^2-\mathcal{K}|\mathcal{K}\ge\mathcal{K}_0^{(T_t)}]}{\mathcal{P}_{\mathcal{K}\le\mathcal{K}_0^{(T_t)}}\,\mathbb{E}[\mathcal{K}|\mathcal{K}\le\mathcal{K}_0^{(T_t)}] + (1-\mathcal{E}^{ST-B})\mathcal{P}_{\mathcal{K}\ge\mathcal{K}_0^{(T_t)}}\,\mathbb{E}[\mathcal{K}|\mathcal{K}\ge\mathcal{K}_0^{(T_t)}]}\frac{\mathbb{E}[\mathcal{K}]}{\mathbb{E}[\mathcal{K}^2-\mathcal{K}]}\mathcal{R}_{\text{eff}}^{ct}, \tag{45}$$

where $\mathcal{R}_{\text{eff}}^{ct} = (1-\zeta\kappa)\mathcal{R}_0$ and $\kappa = \mathcal{R}_0^{sym}/\mathcal{R}_0$ (see §4). The result corresponding to this scenario is shown in Fig. 4D. Accordingly, we predict that $\mathcal{R}_{\text{eff}}^{ST-C} = 1$ can be achieved with 72% app users among smart phone users and 166 tests per 100'000 per day. This is very encouraging, since 72% app users among smart phone users corresponds to only about 50% app users of the whole population. Furthermore, a testing capacity of 166 per 100'000 per day already is available in several developed countries, including Switzerland.

#### 6.4. *Sensitivity Study*

In order to gain further confidence in our STeCC related mitigation scenarios, we conducted studies to investigate the sensitivity of the effective reproduction number for $\mathcal{R}_0 \in \{1.9, 2.9, 3.4\}$. The parameters which we varied are $\gamma \in \{2, 2.5, 3.5\}$ in the exponent of the power-law distribution of the degree of connectivity, the ratio $\eta \in \{0.1, 0.15\}$ of false negatives and the test processing time $\tau_{proc} \in \{0.5, 1.5\}$ (day). Note that our base setting is the combination of $\mathcal{R}_0 = 2.4$, $\gamma = 0.3$, $\eta = 0.05$ and $\tau_{proc} = 1$ (day). Figures S 7 and 8 show the sensitivity of STeCC-B and-C scenarios, respectively, for varying $\mathcal{R}_0$ and $\gamma$ values. Figure S9 depicts the sensitivity of STeCC-C with respect to the fraction of false negatives and test processing time. We observe that for a large range of parameters considered the combination of STeCC and conventional contact tracing leads to stopping the pandemic with realistic app user percentage (i.e., 60% to 85%) and number of tests per day (i.e., 50 to 350 tests per 100'000).

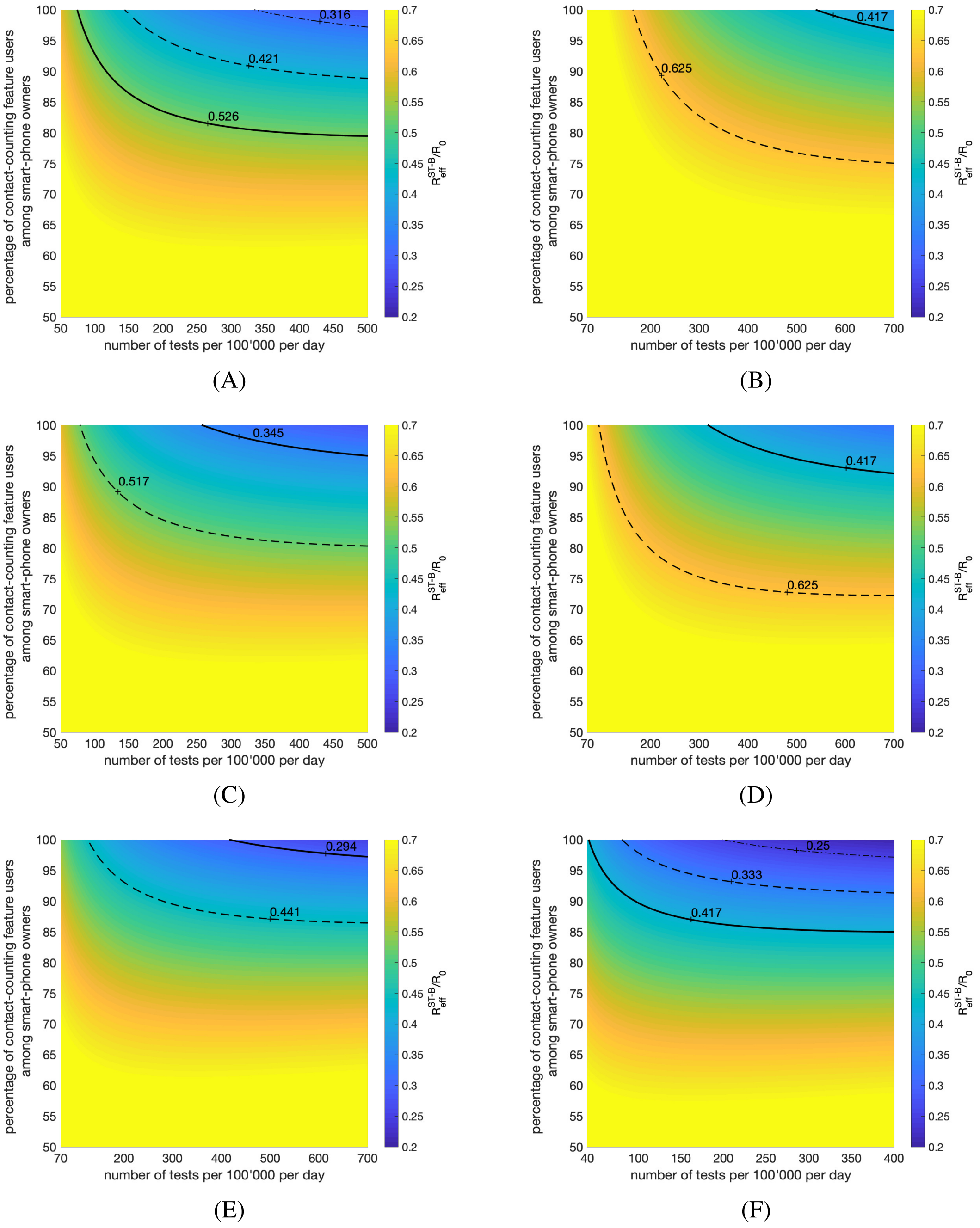


**Fig. S 7.** STeCC plus isolation of contacts with positively tested individuals (STeCC-B): $\mathcal{R}_{\text{eff}}^{ST-B}/\mathcal{R}_0$ as function of number of smart tests per 100′000 per day and the percentage of smart-phone owners who participate in STeCC (scenario B). For ($\gamma = 0.3, k_c = 552$) and (A) $\mathcal{R}_0 = 1.9$, (C) $\mathcal{R}_0 = 2.9$ and (E) $\mathcal{R}_0 = 3.4$; for $\mathcal{R}_0 = 2.4$ and (B) ($\gamma = 0.2, k_c = 227$), (D) ($\gamma = 0.25, k_c = 327$) and (F) ($\gamma = 0.35, k_c = 1325$). A corresponding map with $\mathcal{R}_0 = 2.4$ and $\gamma = 0.3$ is shown in Fig. 4C. The bold lines indicates the combinations for which $\mathcal{R}_{\text{eff}}^{ST-B} = 1$.

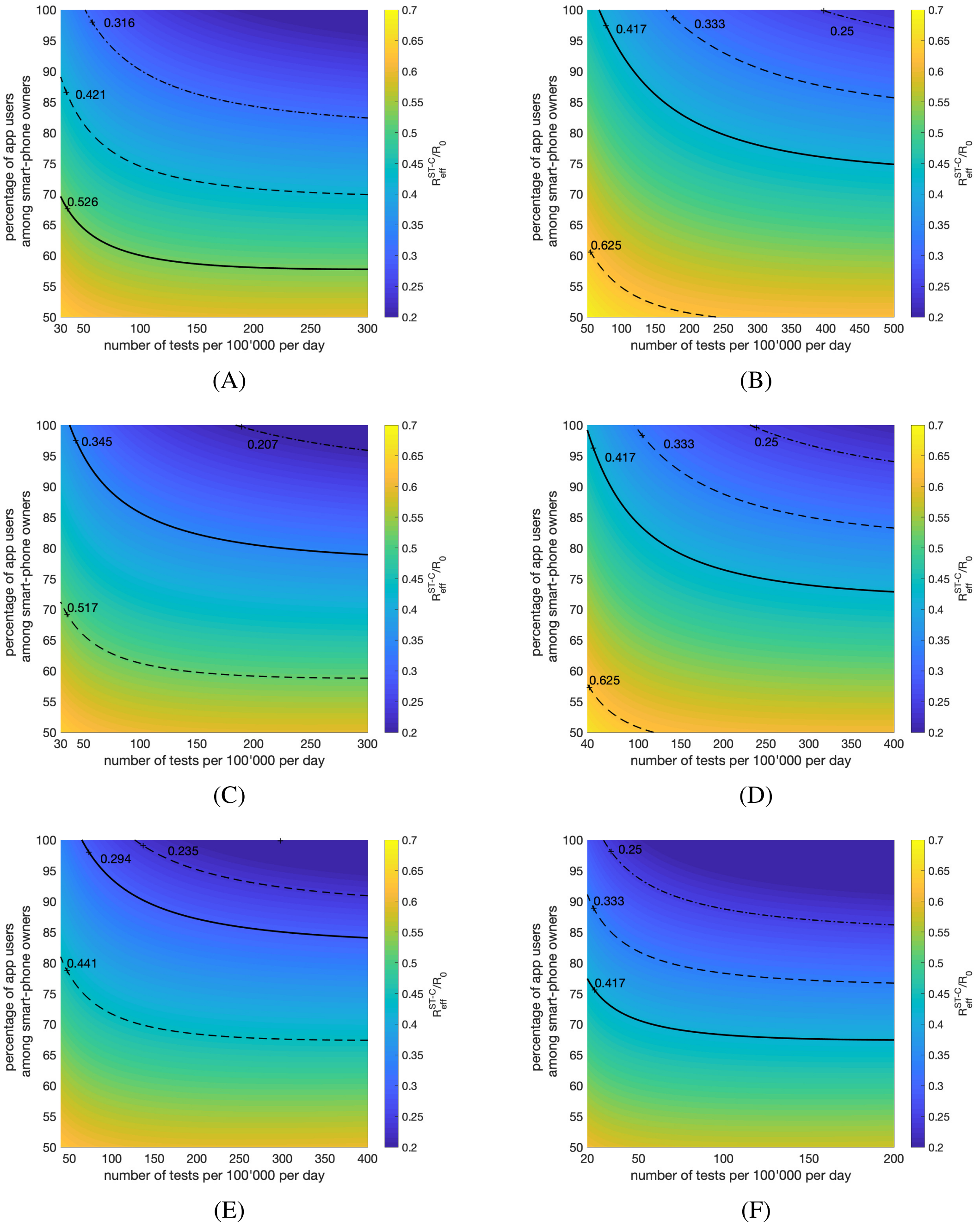


**Fig. S 8.** STeCC plus isolation of contacts with positively tested individuals plus classical contact tracing (STeCC-C): $\mathcal{R}_{\text{eff}}^{ST-C}/\mathcal{R}_0$ as function of number of smart tests per 100′000 per day and the percentage of smart-phone owners who participate in STeCC (scenario C). For ($\gamma = 0.3, k_c = 552$) and (A) $\mathcal{R}_0 = 1.9$, (C) $\mathcal{R}_0 = 2.9$ and (E) $\mathcal{R}_0 = 3.4$; for $\mathcal{R}_0 = 2.4$ and (B) ($\gamma = 0.2, k_c = 227$), (D) ($\gamma = 0.25, k_c = 327$) and (F) ($\gamma = 0.35, k_c = 1325$). A corresponding map with $\mathcal{R}_0 = 2.4$ and $\gamma = 0.3$ is shown in Fig. 4D. The bold lines indicates the combinations for which $\mathcal{R}_{\text{eff}}^{ST-C} = 1$.

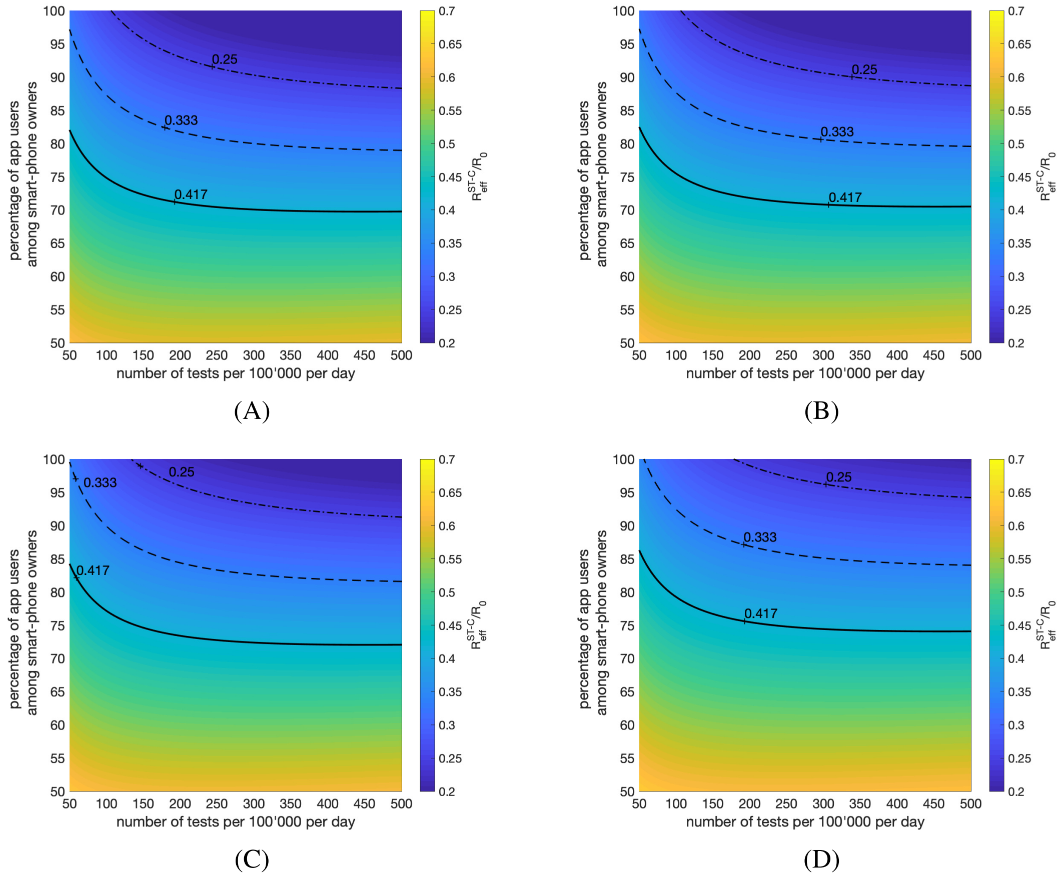


**Fig. S 9.** STeCC plus isolation of contacts with positively tested individuals plus classical contact tracing (STeCC-C): $\mathcal{R}_{\text{eff}}^{ST-C}/\mathcal{R}_0$ as function of number of smart tests per 100′000 per day and the percentage of smart-phone owners who participate in STeCC (scenario C). For ($\eta$ = 0.05) and (A) $\tau_{proc}$ = 0.5 (day) and (C) $\tau_{proc}$ = 1.5 (day); for $\tau_{proc}$ = 1 (day) and (B) $\eta$ = 0.1, (D) $\eta$ = 0.15. A corresponding map with $\eta$ = 0.05 and $\tau_{proc}$ = 1 (day) is shown in Fig. 4D. The bold lines indicates the combinations for which $\mathcal{R}_{\text{eff}}^{ST-C}$ = 1.

### 7. Contact Counting App - One Approach to Ensure Data Privacy

Here we outline one option how an efficient centralized contact counting scheme could be implemented using Fully Homomorphic Encryption (FHE). By centralized we mean that the number of contacts of each user is gathered on a central server every day, which allows to compute the distribution of contacts among the population. Given this distribution allows to determine and broadcast a threshold, beyond which testing is recommended. Subsection 7.1 provides an introduction to FHE, in subsection 7.2 a solution is presented and privacy concerns are discussed in subection 7.3.

#### *7.1. Fully Homomorphic Encryption*

Fully Homomorphic Encryption (FHE) has been the holy grail of cryptographers for a very long time. It is only since 2009, with the seminal work of Gentry [9], that the first concrete and secure construction appeared. However, these initial constructions were far from being practical, and even their mere implementation took few years; see e.g. [10]. Whereas in the beginning performances were catastrophic (keys of few gigabytes and hours to perform operations), recent constructions have proven rather efficient and are now usable in industry. Most notably, the implementation of HElib[2], Seal[3], and TFHE[4] have shown to be of practical value.

A FHE scheme allows to compute on encrypted data. More precisely, let Enc (respectively Dec) be an (asymmetric) encryption algorithm (respectively its corresponding decryption algorithm). These algorithms are non-deterministic, however for the following explanations we will explicitly write the random seed parameter to make the function deterministic (for the same seed value). Let $m_1, \dots m_n \in \mathcal{M}$ be plaintexts (here IDs), and let $f : \mathcal{M}^n \to \mathcal{M}$ be any computational circuit. Let also $c_1, \dots c_n \in C$ be the corresponding ciphertexts, i.e., $c_i = \mathrm{Enc}(m_i)$. Then there exists a circuit $\tilde{f}$ which is efficiently computable such that $\mathrm{Dec}(\tilde{f}(c_1, \dots, c_n)) = f(m_1, \dots, m_n)$, i.e., there is the possibility to apply a function on the plain-texts without decrypting the ciphertexts without knowing their content. It is important to note that most of the practical constructions work at the bit level, which means that only bit operations can be performed on the plaintexts.

#### *7.2. Our Construction*

The goal of our construction is that at the end of each day the central server is aware of the distribution of number of contacts per user, without knowing which user has how many contacts. However, we want to minimize the size of the exchanged messages, i.e., of all the beacons seen that day.

Our idea is to homomorphically update an encrypted counter on each device. This counter will be encrypted with the public key of the server and, thus, only this server will be able to know its value. In more detail:

1. Initially, the central server generates a pair of public/private keys $(\boldsymbol{pk}_c, \boldsymbol{sk}_c)$ for an asymmetric fully homomorphic encryption scheme.
2. Every device will select each day $t$ a new ID that we denote $\mathrm{ID}_t$. This ID is going to be used for contact counting.
3. During each epoch, each device broadcasts a ciphertext of $\mathrm{ID}_t$ with fixed randomness. This means that during one epoch, the same ciphertext is broadcast. More precisely, at epoch $i$ we broadcast the ciphertext $c_i = \mathrm{Enc}_{\boldsymbol{pk}_c}(\mathrm{ID}_t, r_i)$ for some random coins $r_i$. Therefore, between different epochs the ciphertexts will not be linkable, unless one possesses the private key $\boldsymbol{sk}_c$.

[2] `https://github.com/shaih/HElib`
[3] `https://www.microsoft.com/en-us/research/project/microsoft-seal/`
[4] `https://tfhe.github.io/tfhe/`

4. Now we deal with the contact counter ctr. Every day this counter is set to zero; its encription thus is $ctr = \text{Enc}_{\boldsymbol{pk}_c}(0)$[5].
5. Every device will also keep a database of all the beacons seen that day.
6. Upon reception of a beacon $c^*$, which is a ciphertext of an ID, we need to update the counter only if the ID is a new one. We do this by computing the following circuit homomorphically. Let $c_1, \ldots, c_n$ be the beacons that are already stored in the database. We homomorphically compute $d_i = (c^* \oplus c_i)$ for $i \in [1, \ldots n]$, where $\oplus$ is the bitwise XOR. Hence, the plaintext corresponding to $d_i$ is zero, if and only if $c^*$ and $c_i$ come from the same ID.
7. The counter then has to be updated by one only if all the $d_i$ have a corresponding plaintext different from zero. Let $d_{i,1}, \ldots, d_{i,m}$ denote the bits of $d_i$. We will apply the following function $g$ on each $d_i$:

$$g(d_i) = \overline{\overline{d_{i,1}} \& \ldots \& \overline{d_{i,m}}} \ ,$$

   where $\overline{b}$ denotes the NOT operator over the bit $b$. Note that the output of $g$ is the ciphertext of 0, if $d_i$ is the ciphertext of zero, and Dec($g$)=1 otherwise. Hence, we then homomorphically compute ctr=ctr+$g(d_1) \& g(d_2) \& \ldots \& g(d_n)$, where, & denotes the AND operator.
8. At the end of the day this encrypted counter is sent to the central server.

Note that the homomorphic computations can be computationally expensive, which might be problematic on devices that run on a battery (e.g. a mobile phone). A solution to this problem is to delay these homomorphic computations as much as possible (e.g. waiting for the phone to get plugged in). Further, one may reduce the frequency at which beacons are broadcast, thus reducing the number of beacons on which homomorphic computations have to be performed.

Note that minimal communication with the server is required, i.e., only one message has to be sent each day (the encrypted counter).

#### *7.3. Privacy Discussion*

Our scheme has the important advantage of not letting users know whether different beacons at different epochs belong to the same person. Indeed, the properties of the non-deterministic FHE scheme ensures that two ciphertexts of the same plaintext, but with different random coins, are not relatable (IND-CPA security). However, since the central authority knows the associated private key, it could decrypt beacons on the fly and therefore could use this information to track/follow users, i.e. the central authority must be trusted. Attacks that are possible in the DP-3T setting also exist here, e.g. replay and Sybil attacks.

[5] Note that since our scheme operates at bit level, one needs to encode this for many bits.

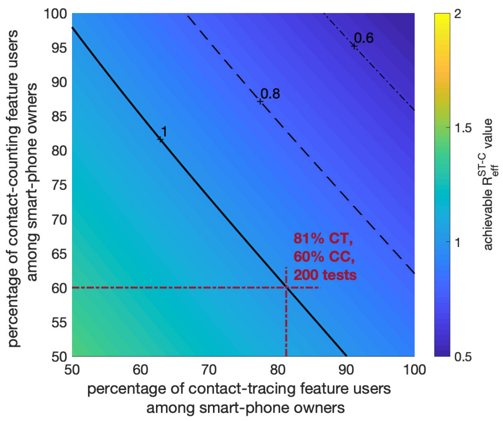


**Fig. S 10.** STeCC-C scenario with different mix of participants: Achievable $\mathcal{R}_{\text{eff}}^{ST-C}$ for 200 tests per 100'000 per day.

Since some users may have concerns with a centralized scheme, we also study how different mixes of contact counting and contact tracing users would affect the performance of STeCC-C, as shown in Fig. 10.

### 8. Model Implementation

The dynamic model was implemented with Maple 2018. The calculations for mass testing, contact tracing and smart testing were implemented with MATLAB and the Statistics Toolbox Release 2018b. The corresponding codes are available on GitHub server via `https://github.com/gorjih2/STeCC_preliminary`.

[7] Luca Ferretti, Chris Wymant, Michelle Kendall, Lele Zhao, Anel Nurtay, Lucie Abeler-Dörner, Michael Parker, David Bonsall, and Christophe Fraser. Quantifying sars-cov-2 transmission suggests epidemic control with digital contact tracing. *Science*, 2020.

[8] Christophe Fraser, Steven Riley, Roy M Anderson, and Neil M Ferguson. Factors that make an infectious disease outbreak controllable. *Proceedings of the National Academy of Sciences*, 101(16):6146–6151, 2004.

[9] Craig Gentry. Fully homomorphic encryption using ideal lattices. In *Proceedings of the 41st annual ACM symposium on Symposium on theory of computing - STOC 09*. ACM Press, 2009. doi: 10.1145/1536414.1536440. URL `https://doi.org/10.1145/1536414.1536440`.

[10] Craig Gentry and Shai Halevi. Implementing gentry's fully-homomorphic encryption scheme. In *Advances in Cryptology - EUROCRYPT 2011 - 30th Annual Inter- national Conference on the Theory and Applications of Cryptographic Techniques - Lecture Notes in Computer Science*. Springer, 2011.

[11] Daniel F Gudbjartsson, Agnar Helgason, Hakon Jonsson, Olafur T Magnusson, Pall Melsted, Gudmundur L Norddahl, Jona Saemundsdottir, Asgeir Sigurdsson, Patrick Sulem, Arna B Agustsdottir, et al. Spread of sars-cov-2 in the icelandic population. *New England Journal of Medicine*, 2020.

[12] Robert Hinch, Will Probert, Anel Nurtay, Michelle Kendall, Chris Wymant, Matthew Hall, Katrina Lythgoe, Ana Bulas Cruz, Lele Zhao, Andrea Stewart, Luca Ferretti, Michael Parker, Ares Meroueh, Bryn Mathias, Scott Stevenson, Daniel Montero, James Warren, Nicole K Mather, Anthony Finkelstein, Lucie, Abeler-Dörner, David

Bonsall, and Christophe Fraser. Effective configurations of a digital contact tracing app: A report to NHSX, April 2020. URL `https://045.medsci.ox.ac.uk/files/files/report-effective-app-configurations.pdf`.

[13] Ayaan Hossain, C. Alexander Reis, Sarthok Rahman, and M. Howard Salis. A massively parallel covid-19 diagnostic assay for simultaneous testing of 19200 patient samples, 2020.

[14] Enrico Lavezzo, Elisa Franchin, Constanze Ciavarella, Gina Cuomo-Dannenburg, Luisa Barzon, Claudia Del Vecchio, Lucia Rossi, Riccardo Manganelli, Arianna Loregian, Nicolò Navarin, et al. Suppression of covid-19 outbreak in the municipality of vo, italy. *medRxiv*, 2020.

[15] Ruiyun Li, Sen Pei, Bin Chen, Yimeng Song, Tao Zhang, Wan Yang, and Jeffrey Shaman. Substantial undocumented infection facilitates the rapid dissemination of novel coronavirus (sars-cov2). *Science*, 2020.

[16] Ying Liu, Albert A Gayle, Annelies Wilder-Smith, and Joacim Rocklöv. The reproductive number of covid-19 is higher compared to sars coronavirus. *Journal of travel medicine*, 2020.

[17] Zhonghua Liu, Xing Bing, and Xue Za Zhi. The epidemiological characteristics of an outbreak of 2019 novel coronavirus diseases (covid-19) in china. *Novel Coronavirus Pneumonia Emergency Response Epidemiology Team*, 41(2):145, 2020.

[18] Robert M May and Alun L Lloyd. Infection dynamics on scale-free networks. *Physical Review E*, 64(6):066112, 2001.

[19] Kenneth McIntosh, Martin S Hirsch, and Allyson Bloom. Coronavirus disease 2019 (covid-19). *UpToDate*, 5, 2020.

[20] Ke Men, Xia Wang, Yihao Li, Guangwei Zhang, Jingjing Hu, Yanyan Gao, and Henry Han. Estimate the incubation period of coronavirus 2019 (covid-19). *medRxiv*, 2020.

[21] Lauren Ancel Meyers, Babak Pourbohloul, Mark EJ Newman, Danuta M Skowronski, and Robert C Brunham. Network theory and sars: predicting outbreak diversity. *Journal of theoretical biology*, 232(1):71–81, 2005.

[22] Kenji Mizumoto and Gerardo Chowell. Estimating risk for death from 2019 novel coronavirus disease, china, january-february 2020. *Emerging infectious diseases*, 26(6), 2020.

[23] Joël Mossong, Niel Hens, Mark Jit, Philippe Beutels, Kari Auranen, Rafael Mikolajczyk, Marco Massari, Stefania Salmaso, Gianpaolo Scalia Tomba, Jacco Wallinga, et al. Social contacts and mixing patterns relevant to the spread of infectious diseases. *PLoS medicine*, 5(3), 2008.

[24] Mark Newman. *Networks*. Oxford university press, 2018.
